# Prompting is All You Need: How to Make LLMs More Helpful for Clinical Decision Support

**DOI:** 10.64898/2026.02.12.26346005

**Authors:** Braydon Dymm, Daniel M. Goldenholz

## Abstract

**Importance:** Large language models (LLMs) offer potential decision support, but their accuracy varies. Prompt engineering can generally enhance LLM behavior in a clinical context, yet best practices have yet to be formally explored in realistic neurology settings.

**Objective:** To evaluate the impact of structured prompting versus simple prompting on the performance of six LLMs (three closed-source: OpenAI GPT-4o, OpenAI o3, OpenAI GPT-5.2 Thinking; three open-source: Meta Llama-4-Scout-17B-16E-Instruct, Llama-3.3-70B-Instruct-Turbo, and the reasoning model R1-1776) for thrombolytic clinical decision support (CDS) in acute stroke.

**Design:** Models responded to three novel ischemic stroke vignettes using either a simple question (“Should this patient be offered thrombolytics?”) or a five-step structured prompt (CARDS) guiding information extraction, timing analysis, contraindication checking, decision process explanation, and risk-benefit discussion. Outputs were assessed across seven domains: guideline adherence, unsafe recommendations, risk recognition, guideline grading accuracy, inclusion of conversational explanation, clarity, and overall helpfulness.

**Results:** Structured prompts significantly enhanced performance across most domains, with varying effects between model families. For some closed-source models (GPT-4o, o3), prompts structured in the CARDS style improved guideline adherence from 83.3% to 100%, eliminated unsafe recommendations (16.7% to 0%), and increased specific guideline grading accuracy from 0% to 100%. The closed-source reasoning model GPT-5.2 Thinking similarly achieved 100% adherence, 0% unsafe recommendations, and 100% grading accuracy with structured prompts, while also maintaining perfect safety and risk recognition under simple prompting. Similarly, the open-source reasoning model R1-1776 achieved these top-tier outcomes (100% adherence, 0% unsafe, 100% grading, 100% conversation) when structured prompts were applied, with grading and conversation improving from 0%. In contrast, other open-source models (Llama-4-Scout, Llama-3.3-70B) showed more modest gains: risk recognition improved (83.3% to 100%) and guideline grading accuracy increased (0% to 66.7%), while guideline adherence (66.7%) and unsafe recommendations (33.3%) persisted. Overall, structured prompting yielded the largest improvements in guideline grading accuracy and conversational reasoning across multiple models.

**Conclusion:** Structured prompting substantially enhances LLM performance for acute stroke thrombolysis CDS. Notably, some models, including the proprietary GPT-4o, o3, and GPT-5.2 Thinking, and the open-source reasoning model R1-1776, achieved excellent safety and adherence with structured prompts. For clinical deployment of any LLM, structured prompts are crucial, and vigilant human oversight remains essential.

## Introduction

Large language models (LLMs) have emerged as potential decision□support tools capable of parsing free□text clinical data, for example identifying thrombolysis contraindications.^1^ Early evaluations, however, reveal variable accuracy and occasional unsafe advice when models are queried with simple, unstructured prompts.^2^ Recent work in medical question□answering demonstrates that carefully engineered prompts—explicit instructions that force the model to extract facts, reason step□by□step, and cite guidelines—can boost performance more than additional fine□tuning.^3^ Clinician-focused guidance on prompt engineering for clinical practice has also emerged.^4^ Yet few studies have compared the impact of prompt structure across proprietary and open□source LLMs while focusing on high□stakes, time□critical interventions such as tPA decisions.

Decisions regarding tPA are central to the management of ischemic stroke. This condition remains a leading cause of death and disability worldwide, and its absolute burden continues to rise despite decades of prevention efforts.^5^ Intravenous tissue□type plasminogen activator (tPA) delivered within the first hours after symptom onset improves functional outcome but carries bleeding risk.^6^ Achieving timely, guideline□concordant thrombolysis is challenging; real□world registries show that emergency department and workflow factors frequently delay door□to□needle time and introduce protocol deviations.^7^

We therefore assessed six contemporary LLMs: three closed (OpenAI GPT□4o, OpenAI□o3, and OpenAI GPT□5.2 Thinking) and three open (Meta□Llama□4□Scout□17B□16E□Instruct, Llama□3.3□70B□Instruct□Turbo, and Perplexity R1-1776) on standardized acute stroke vignettes. By contrasting a minimalist question prompt with a five□step structured rubric, we aimed to quantify how prompt engineering influences safety, guideline adherence, and explanatory quality in thrombolytic decision support. This introduction sets the stage for a systematic evaluation designed to inform both future model development and pragmatic clinical deployment.

## Methods

Six large language models, 3 closed (OpenAI GPT□4o, OpenAI o3, OpenAI GPT□5.2 Thinking) and 3 open (Meta□Llama□4□Scout□17B□16E□Instruct, Meta Llama□3.3□70B□Instruct□Turbo, and Perplexity R1-1776), were evaluated. Of note, R1-1776 is a model derived from DeepSeek-R1, with authoritarian censorship removed using fine-tuning at Perplexity Labs. Each model responded to three novel, synthetic ischemic stroke vignettes using two prompt structures (Appendix A). The vignettes are not based on actual patients; they were constructed by the authors for the purpose of this evaluation. The simple prompt contained only the vignette and the question, “Should this patient be offered thrombolytics?”

The structured prompt followed a simple, easy-to-remember framework called CARDS, which stands for Context, Aims, Relevant details, Design, and Source (Figure 1). The prompt included clear instructions to guide the model through five key steps: extracting critical information, analyzing timing, checking for contraindications, explaining the decision process, and discussing the risks and benefits. All queries for OpenAI’s 4o and o3 models and Llama models were sent through the respective Application Programming Interfaces (APIs) on 6□May□2025 with default parameters and no system prompts. Queries for GPT-5.2 Thinking Low were sent on 12 February 2026 through AvoMD with default parameters and no system prompts. The API provider for the closed models was AvoMD, and the provider for the Llama models was Together AI. Queries for R1-1776 were sent on 23 May 2025 through provider Perplexity Labs.

**Figure 1.**
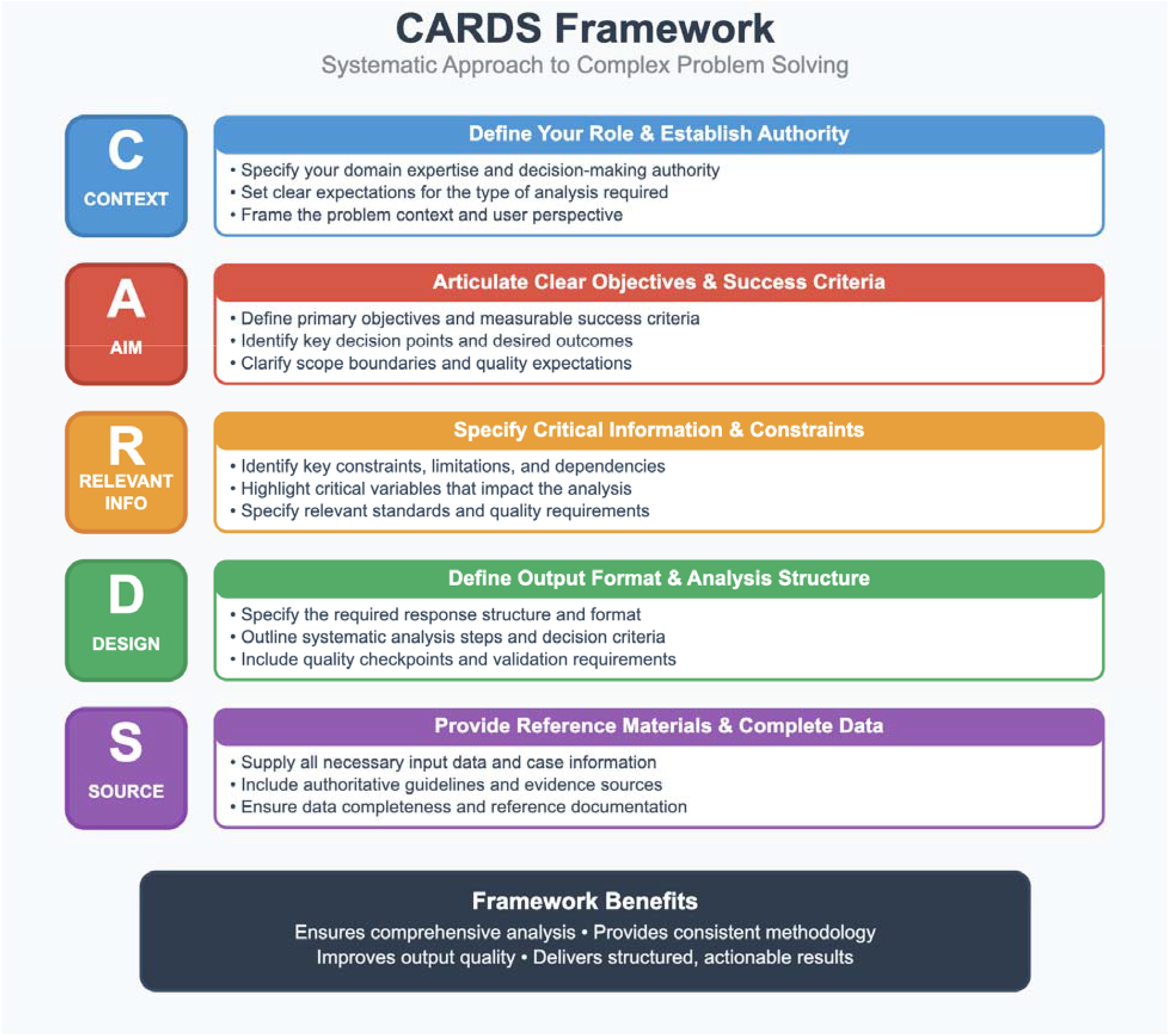
CARDS Framework.

Seven performance domains were manually rated by a board-certified stroke neurologist: guideline adherence (yes/no), safety of recommendations (yes/no), recognition of key risks (yes/no), accuracy of specific guideline grading (yes/no), inclusion of a conversational explanation (yes/no), as well as clarity and overall helpfulness on a five□point scale from very poor (1) to very good (5).

### Standard Protocol Approvals, Registrations, and Patient Consents

This study did not involve human participants; it used only synthetic, fictional clinical vignettes constructed for evaluation. No experiments using human participants or live vertebrates were performed. No institutional or regional ethical standards committee approval was required for this work. No written informed consent was required because no human participants were enrolled. No photographs, videos, or other identifiable information pertaining to real individuals are included; consent to disclose was therefore not applicable. This was not a clinical trial; no trial registration or protocol submission is reported.

## Results

Structured prompts enhanced performance across six of the seven evaluated domains, with varying degrees of improvement between LLMs (Table 1). We observed distinct patterns of improvement between closed-source commercial models and open-source models.

**Table 1.**
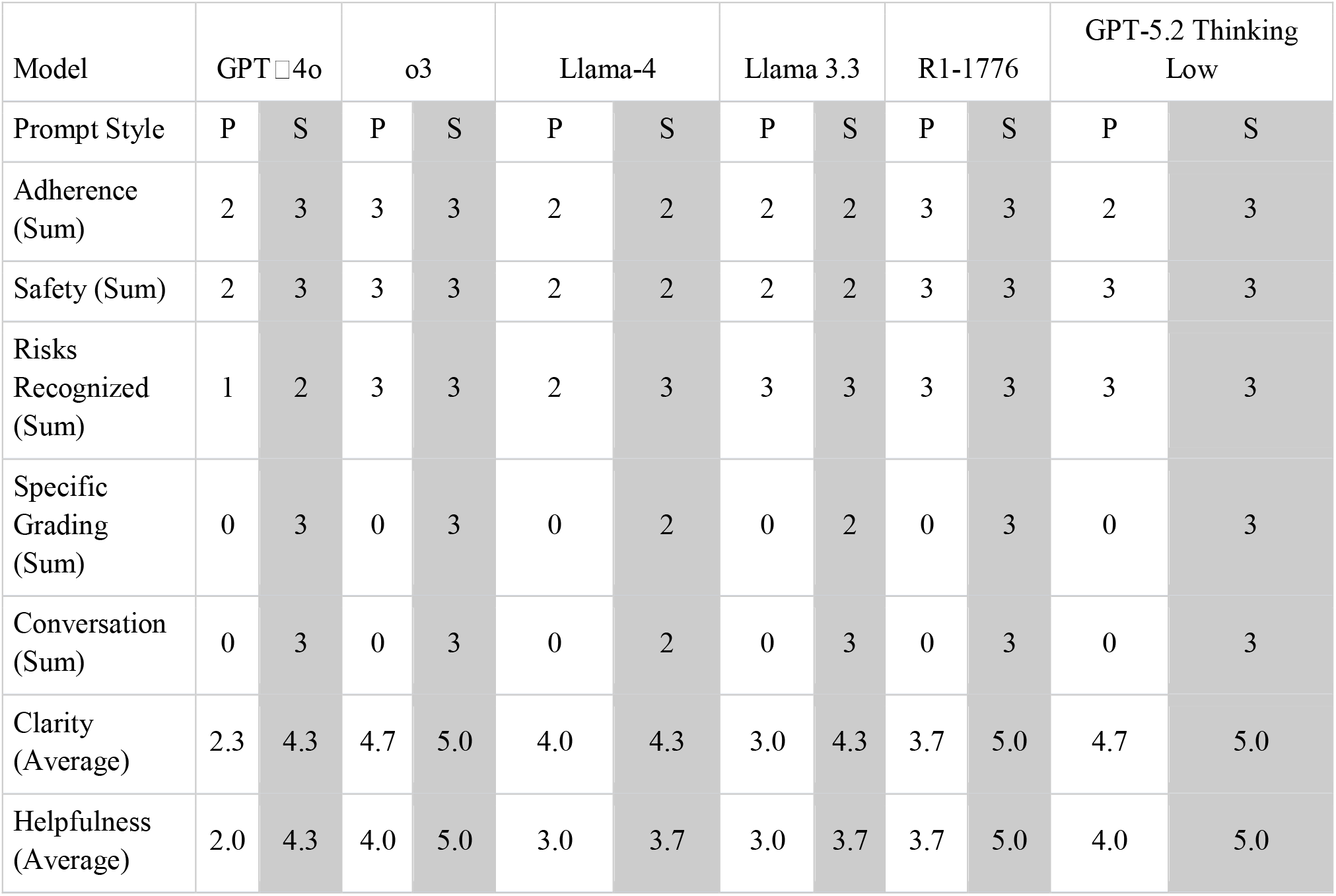
Summary results of model responses. P = Simple. S = Structured.

### Closed models (GPT□4o and o3)

Closed-source models demonstrated substantial improvements across multiple domains when using structured prompts (Table 1). Outputs from the 3 cases in both models are considered here. Adherence to clinical guidelines increased from 83.3% (5 of 6 outputs) to 100% (6 of 6 outputs). Notably, unsafe recommendations, which appeared in 16.7% (1 of 6) of simple outputs, were eliminated in structured prompt responses. Risk recognition improved from 66.7% (4 of 6) to 83.3% (5 of 6). The 3 risk failures were each from 4o which failed to recognize at least one specific risk in each case.

The most dramatic improvements were observed in specific guideline grading accuracy, which increased from 0% (0 of 6) with simple prompts to 100% (6 of 6) with structured prompts.Similarly, conversational explanations, entirely absent in simple outputs, appeared in 100% (6 of 6) of structured outputs. Response clarity improved significantly, shifting from a mixed distribution (50% good, 16.7% neutral, 33.3% poor) to uniformly good ratings (100%).

### Closed reasoning model GPT-5.2 Thinking

GPT-5.2 Thinking demonstrated strong baselines even under simple prompting: safety was perfect (0 of 3 unsafe) and risk recognition was 100% (3 of 3) (Table 1). Guideline adherence improved from 66.7% (2 of 3) with simple prompts to 100% (3 of 3) with structured prompts; the sole simple-prompt adherence failure occurred in Case C, where the model issued a flat refusal based on the prior stroke within 3 months rather than recommending shared decision-making.

Guideline grading accuracy increased from 0% (0 of 3) to 100% (3 of 3), and conversational explanations increased from 0% to 100% with structured prompts. Response clarity improved from 4.7 to 5.0, and mean helpfulness increased from 4.0 to 5.0, with all three structured outputs achieving “Very Good” ratings.

### Non-reasoning Open models (Llama□4□Scout□17B□16E□Instruct, and Llama□3.3□70B□Instruct□Turbo)

Open-source models showed more modest but still meaningful improvements with structured prompts (Table 1). Guideline adherence remained unchanged at 66.7% (4 of 6 outputs).Similarly, unsafe recommendations persisted at the same rate (33.3%, 2 of 6 outputs) despite structured prompting.

Risk recognition improved from 83.3% (5 of 6) to 100% (6 of 6) with structured prompts.Guideline grading accuracy increased from 0% (0 of 6) to 66.7% (4 of 6). The presence of conversational explanations increased substantially from 0% to 83.3% (5 of 6 outputs).

Response clarity improved from a mixed distribution (66.7% good, 16.7% neutral, 16.7% poor) to uniformly good ratings (100%). Mean helpfulness ratings increased moderately from 3.0 to 3.7, with structured prompting adding two “Very Good” ratings not present in simple outputs.

*Reasoning open model R1-1776*. For R1-1776, guideline adherence remained unchanged at 100% (3 of 3 outputs) with structured prompts (Table 1). Similarly, unsafe recommendations remained at 0% (0 of 3 outputs), meaning all outputs were safe. Risk recognition also remained consistently high at 100% (3 of 3 outputs) for both simple and structured prompting.

However, significant improvements were observed with structured prompts in other areas: guideline grading accuracy increased from 0% (0 of 3) to 100% (3 of 3), and the presence of conversational explanations increased substantially from 0% (0 of 3) to 100% (3 of 3 outputs). Response clarity improved from a mixed distribution with simple prompts (66.7% ‘Good’ [2 of 3], 33.3% ‘Neutral’ [1 of 3]) to uniformly ‘Very Good’ ratings (100%, 3 of 3) with structured prompts. Mean helpfulness ratings for R1-1776 increased from 3.7 with simple prompts to 5.0 with structured prompts, the latter adding three ‘Very Good’ ratings not present in its simple outputs. The examples of risk/benefit discussions contributed to the response’s helpfulness (Figure 2).

**Figure 2.**
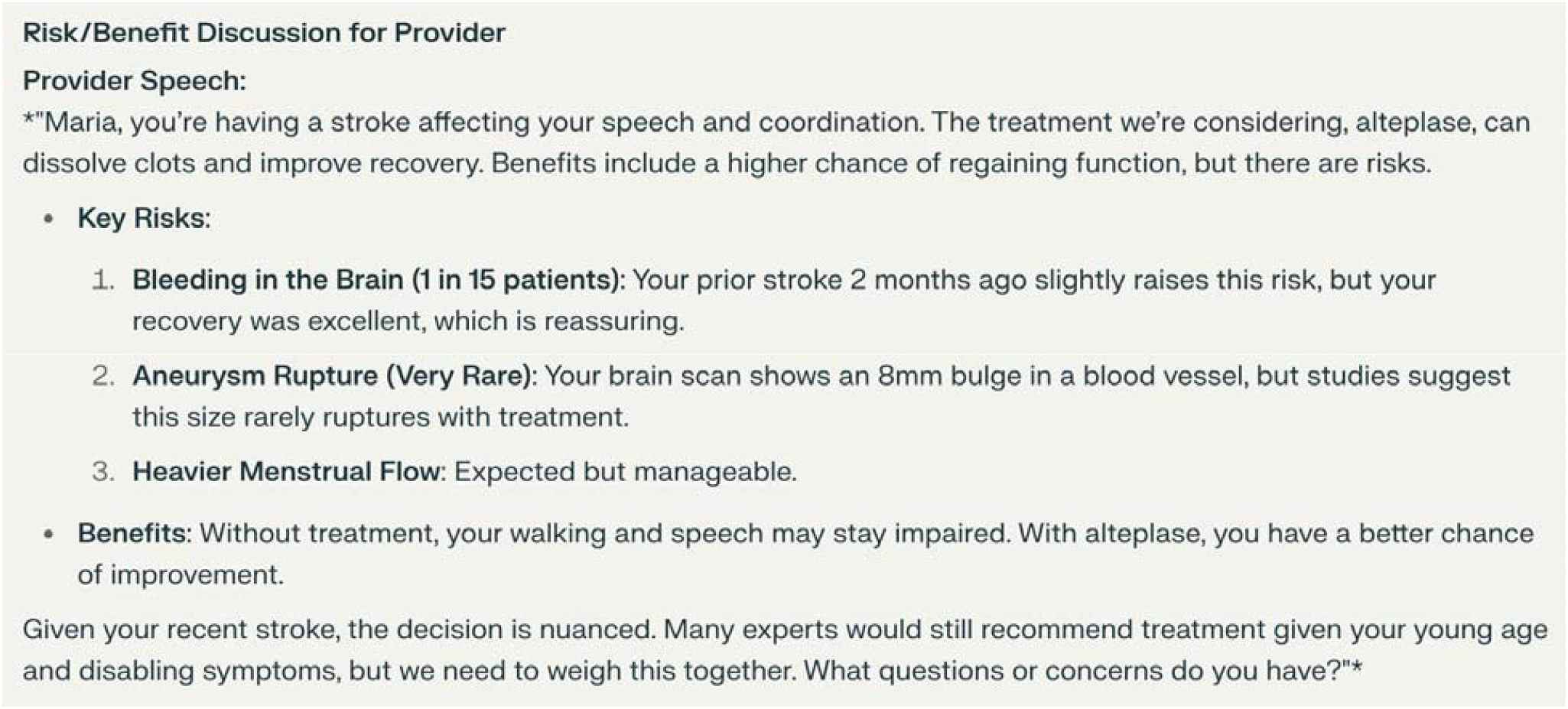
Example of Risk/Benefit Discussion by R1-1776 on Case C.

*Overall*. When examining results across all models, the structured prompting approach produced the largest absolute gains in guideline grading accuracy (from 0 to 16 correct outputs across all model outputs) and in the addition of conversational reasoning (from 0 to 17 outputs). Full results are shown in Table 2 and Appendix B. Safety improvements and adherence gains were more consistently observed in closed-source models, suggesting these models may be more responsive to structured guidance for safety-critical aspects.

**Table 2.**
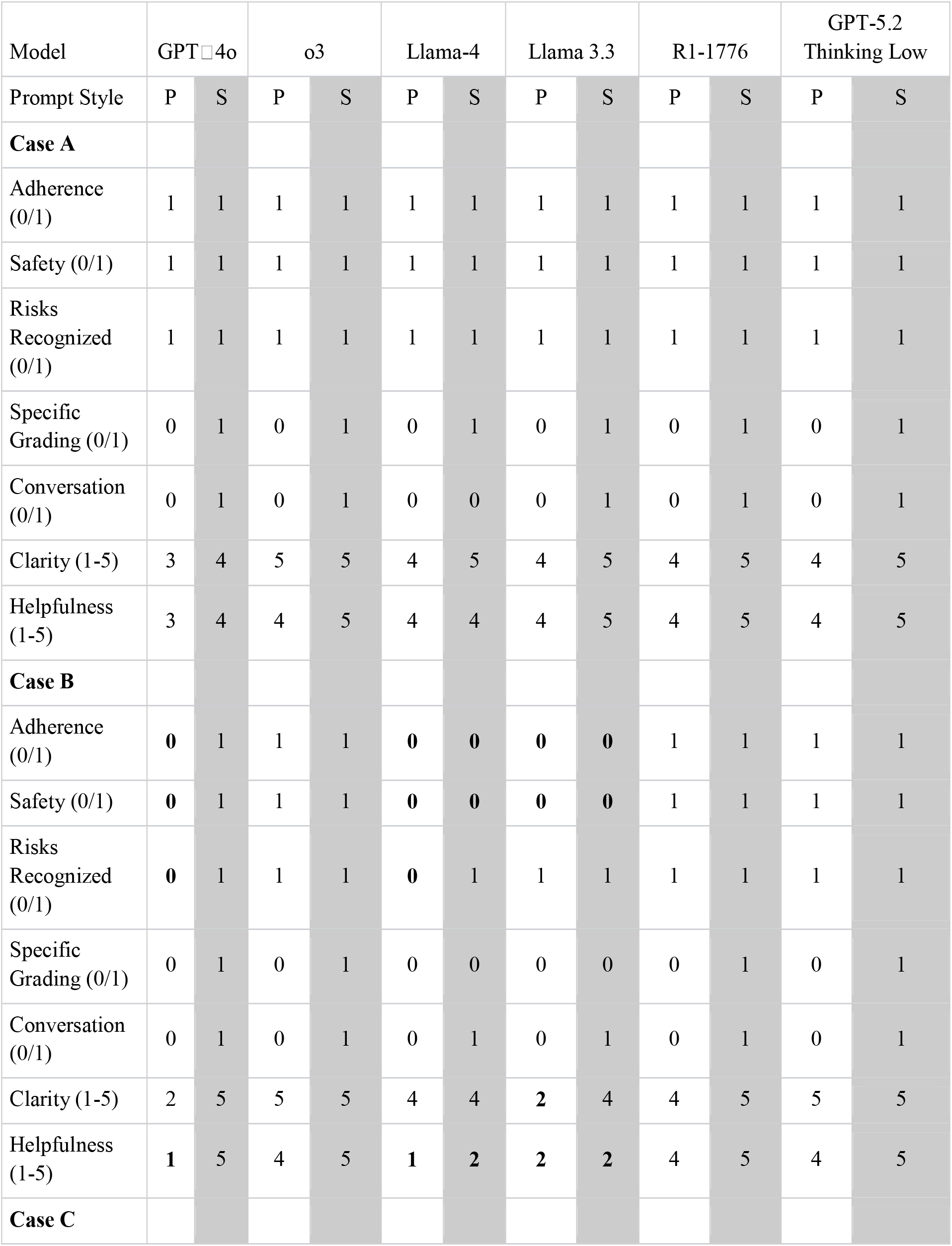
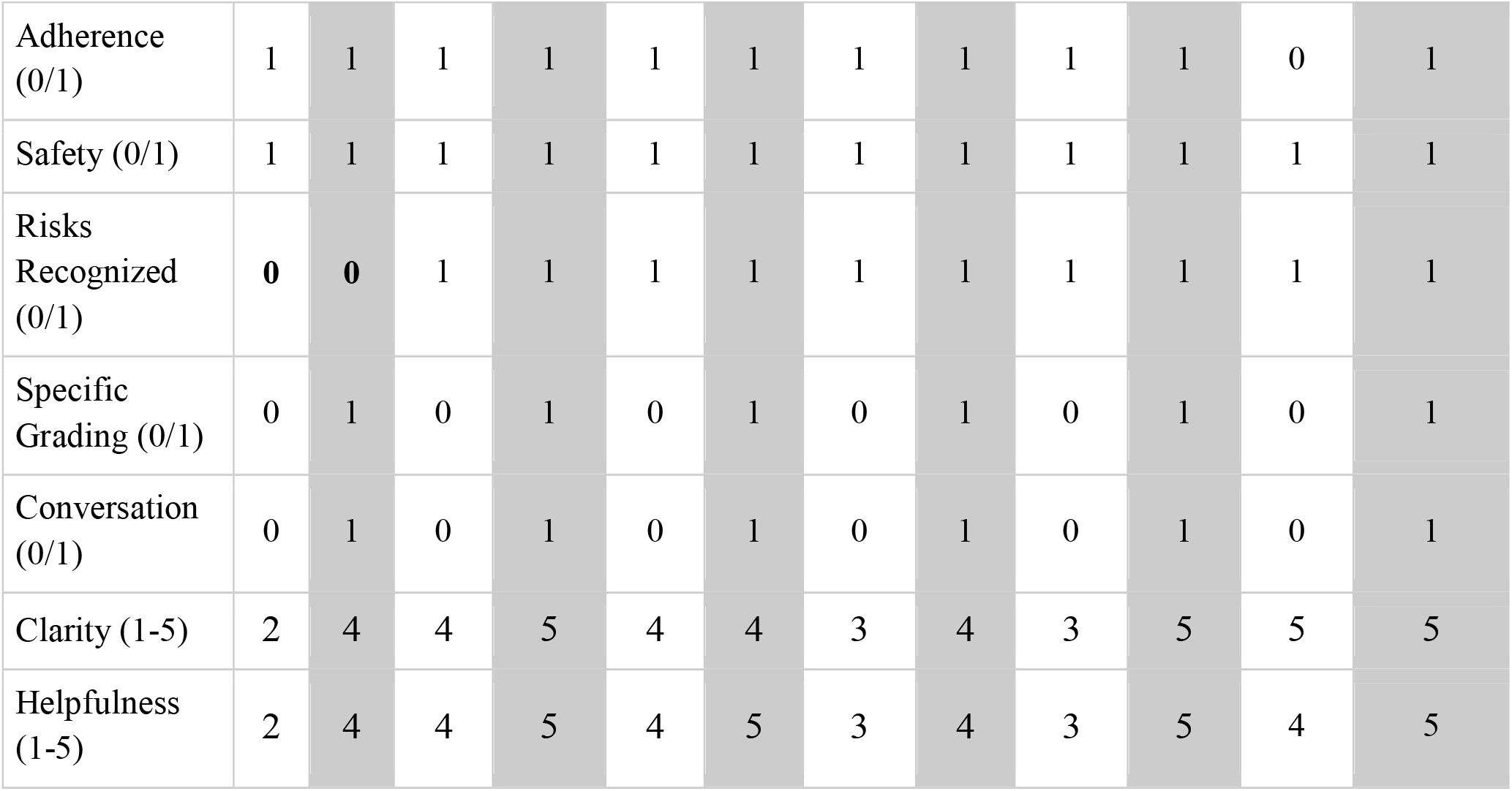
Full results of model responses. P = Simple. S = Structured.

## Discussion

### Main findings and interpretation

The five□step structured prompt markedly sharpened thrombolytic clinical decision support, but the size and nature of the gain depended on the model. Proprietary engines (GPT□4o, o3, and GPT□5.2 Thinking) handled simple prompts reasonably well and, with structured prompting, achieved 100% guideline adherence, eliminated a prior unsafe suggestion, and converted every answer into a clear, conversational explanation with correct guideline grading. For these models, the most significant improvements were in guideline grading and explanatory depth, often moving from low or 0% to 100%.

The performance of open-source models varied. The open-source reasoning model, R1-1776, demonstrated exceptional improvement with structured prompts, achieving 100% in guideline adherence, safety (no unsafe recommendations), risk recognition, specific guideline grading, and the inclusion of conversational explanations. This performance was notably strong, with guideline grading and conversational explanations improving from 0% with simple prompts to 100% with structured prompts. It is worthwhile emphasizing that R1-1776, alongside the proprietary models o3 and GPT-5.2 Thinking (also characterized as reasoning models), showcased how even models designed for reasoning benefit significantly from structured prompting. In contrast, other open□source models (Llama□4□Scout□17B□16E□Instruct and Llama□3.3□70B□Instruct□Turbo) also showed improvements with structured prompts, such as risk recognition climbing to 100% and guideline□grading precision reaching 66.7%. However, these Llama models remained flat on adherence (66.7%) and retained two unsafe recommendations even with structured prompts. The disparities observed, particularly between R1-1776 and other open-source models, likely reflect differences in training data scope, model architecture, reasoning capabilities, alignment pipelines, and built□in medical safeguards, not solely prompt design.

## Relationship to prior work

Prompt□structure effects parallel recent bench studies. Sonoda et□al. showed that a structured clinical□reasoning prompt boosted diagnostic accuracy of an LLM by 18□percent on radiology vignettes.^8^ Singhal and colleagues reported that LLMs harbor latent clinical knowledge that is unlocked by instruction tuning and careful prompting.^9^ Our data extend these observations to acute□stroke management. While structured prompting yielded significant improvements across models, the specific areas and magnitude of gains varied. For instance, all three proprietary models and the open-source reasoning model R1-1776 demonstrated substantial enhancements in guideline grading and explanatory depth, often from 0% to 100% with structured prompting. Other open-source models, like the Llama family, also benefited, though to a lesser extent in certain metrics such as guideline adherence and safety when starting from weaker baselines.

### Limitations

Generalizability is limited by three vignettes, six well-known LLMs, a single rater, default temperature settings, and a single API snapshot. Alternative LLMs, rubrics or parameter sweeps might yield different results. Observed differences in performance between models could reflect factors such as model size, recency, specific training datasets (including the extent of medical data), or computational resources used for tuning, rather than solely their proprietary/open-source status or general architecture.

### Clinical and research implications

For near□term use, clinicians should pair LLMs with structured prompts that require extraction, timing analysis, contraindication checks, and explicit risk–benefit reflection. The closed□source 4o, o3, and GPT□5.2 Thinking models delivered the most reliable safety and adherence, perhaps making them preferable in high□stakes settings.

Some open□source LLMs may need supervised fine□tuning against thrombolytic guidelines or retrieval□augmented generation from verified sources before clinical deployment.^10^ The open-source thinking model R1-1776 with appropriate prompting (or models of similar capability) may represent the ideal balance of privacy preserving (through local model runs) and strict adherence to the criteria recommended here.

### Future directions

Larger vignette sets, multiple expert adjudicators, more frontier LLMs, and head□to□head tests of alternative prompt designs are needed. Longitudinal tracking across model version releases will clarify whether intrinsic safety gaps are shrinking. Experiments that fine□tune open models on stroke□care corpora could reveal whether the observed adherence deficit is modifiable. Until such data emerge showing perfect and safe results, structured prompting plus vigilant human oversight remains the safest path for LLM□assisted thrombolytic decision support.

## Conclusion

Our systematic evaluation demonstrates that structured prompting significantly enhances LLM performance in acute stroke thrombolysis clinical decision support. The five-step rubric (information extraction, time analysis, contraindication check, decision process, and risk-benefit discussion) consistently improved model outputs across multiple domains. The extent of these improvements and final performance levels varied across the models tested. For instance, proprietary models (GPT-4o, o3, and GPT-5.2 Thinking) and the open-source reasoning model R1-1776 achieved perfect (100%) guideline adherence and eliminated unsafe recommendations with structured prompts. In contrast, other open-source models (Llama-4-Scout and Llama-3.3-70B) showed improvements in risk recognition and explanatory quality with structured prompting but maintained gaps in adherence and safety. These findings suggest that while prompt engineering is a powerful tool, its ability to elicit optimal performance may depend on specific model architecture, training, and alignment, with some open-source models demonstrating capabilities comparable to proprietary ones when appropriately prompted.

For clinical implementation, we recommend using structured prompts that enforce systematic reasoning steps. Models that demonstrated superior safety and adherence profiles with such prompting, including the proprietary GPT-4o, o3, and GPT-5.2 Thinking, as well as the open-source R1-1776, are preferable starting points. However, human oversight remains essential for all LLM-assisted clinical decision-making, particularly when deploying any model in high-stakes scenarios until extensive validation is complete. Future research should include larger vignette sets, multiple expert raters, and exploration of model fine-tuning specific to stroke care protocols, which could further enhance models like the Llama family that did not reach optimal performance with prompting alone. As LLMs continue to evolve, the combination of careful prompt engineering and rigorous model selection, considering high-performing open-source options alongside proprietary ones, offers a promising approach to enhance stroke care decision support, potentially improving guideline adherence and reducing treatment delays in this time-critical clinical scenario.

## Data Availability

All data produced in the present study are available upon reasonable request to the authors

## Acknowledgments

The authors would like to thank Valerie Sharf, MD, and Kirvia Williams, DO, for their review of the cases presented in this manuscript.

## Author Contributions

BD and DMG contributed to the conception and design of the study; BD contributed to the acquisition and analysis of data; BD and DMG contributed to drafting the text or preparing the figures.

### Funding

DMG received funding from NIH K23NS124656.

### Data Availability

The data that support the findings of this study are available from the corresponding author upon reasonable request.

