## Appendix A for "Prompting is All You Need: How to Make LLMs More Helpful for Clinical Decision Support"

1. Cases
A) **Straightforward Case Vignette**

Tina Patel is a 55-year-old woman with well-controlled hypertension who presented to the emergency department 75 minutes after the sudden onset of right facial droop and right arm weakness. Her NIHSS score is 9. Her blood glucose is 125 mg/dL, and a CT head showed no hemorrhage or frank hypodensity. She has not any history of major surgery, major bleeding, or any cancer. Her blood pressure is currently 190/115 mmHg. Should this patient be offered thrombolytics?

B) **Difficult Case Vignette**

Joseph Mercado is a 78-year-old man with atrial fibrillation (on warfarin) and type 2 diabetes who presents after being found with aphasia and severe left-sided weakness. He was last seen normal 3 hours ago. His NIHSS score is 28. His current INR is 1.8, and his blood pressure is 175/90 mmHg. A CT head shows early subtle ischemic changes but no significant hypodensity or hemorrhage. He has no recent major surgery or major bleeding. He had basal cell carcinoma that was excised last week. Should this patient be offered thrombolytics?

C) **Indeterminate Case Vignette**

Maria Gonzales is a 42-year-old woman, with a history of small lacunar stroke of the left basal ganglia 2 months ago with full recovery, who presents 2 hours after the acute onset of dysarthria, difficulty walking, and left-sided incoordination. Her NIHSS score is 6. She has a known 8 mm unruptured intracranial aneurysm of the anterior communicating artery and is currently menstruating with a normal flow. She reports no history of intracranial hemorrhage related to the prior stroke, no recent major surgery, or cancer. Her blood pressure is currently 140/80 mmHg, and her CT head is negative for hemorrhage. Should this patient be offered thrombolytics?

2. CARDS Prompt

### **Context**

You are a specialized medical AI assistant tasked with analyzing complex stroke patient vignettes. You need to determine whether thrombolytic therapy (tPA/alteplase) should be offered based on clinical guidelines, contraindications, and time-sensitive considerations. Stroke cases often present with nuanced clinical features that require careful consideration of risk-benefit profiles.

### **Aim**

Accurately assess whether a patient with stroke symptoms should be offered thrombolytic therapy by:

1. Systematically identifying relevant clinical information from patient vignettes
2. Applying evidence-based stroke management guidelines
3. Recognizing both absolute and relative contraindications
4. Weighing time-dependent factors critical for treatment decisions
5. Providing a clear recommendation with supporting rationale

### **Relevant Details**

- Your analysis for determining thrombolytic therapy eligibility must be exclusively based on the information contained within the attached patient case vignette and the criteria specified in the provided clinical guidelines. These guidelines come with class of recommendation and level of evidence, which should be explicitly noted when referenced. These are your sole reference sources; no external data or assumptions should be used. The detailed clinical data points and systematic approach outlined in the 'Design' section below must be meticulously applied.

### **Design**

When analyzing a patient vignette, follow this systematic approach:

1. **Extract Critical Information**:
   - Patient demographics (age, sex)
   - Precise time of symptom onset
   - Current time of evaluation
   - Neurological deficits (with NIHSS if provided)
   - Vital signs, especially blood pressure
   - Relevant lab values (platelets, INR, glucose)
   - Imaging findings
   - Medical history and medications
2. **Time Analysis**:
   - Calculate exact time since symptom onset
   - Determine which time window applies (0-3h, 3-4.5h, >4.5h, or wake-up/unknown)
3. **Contraindication Check**:
   - Systematically assess for each absolute contraindication
   - Identify any relative contraindications
   - Note any special circumstances that require additional consideration
   - For each, if there is a Class of Recommendation and Level of Evidence that applies, include it here.
4. **Decision Process**:
   - If within time window and no absolute contraindications that can not be quickly corrected: Recommend thrombolysis
   - If within time window but with relative contraindications or complicated cases with limited data: Requires shared decision making
   - If absolute contraindications present: Do not recommend thrombolysis
5. **Response Format**:
   - Provide a clear "Yes," "No," or "Requires shared decision making with modified risk/benefit discussion" recommendation regarding thrombolytic therapy
   - Support with specific reasoning tied to guidelines
   - If "No," suggest appropriate alternative management
   - Provide a Risk / Benefit discussion speech to be given by the provider to the patient.

For each patient vignette, start by restructuring the information into these categories before forming your conclusion. This ensures no critical details are missed.

### **Source**

Case: <<case goes here>>

Guidelines: <<guidelines go here>>

Appendix B

1. OpenAI 4o (Simple Prompting) - Case A

| **Parameter** | **Details** |
| --- | --- |
| Age | 55 years old |
| Condition | Well-controlled hypertension |
| Symptoms | Right facial droop, right arm weakness |
| Time since onset | 75 minutes |
| NIHSS Score | 9 |
| Blood Glucose | 125 mg/dL |
| CT Head | No hemorrhage or frank hypodensity |
| Medical History | No major surgery, major bleeding, or cancer |
| Current Blood Pressure | 190/115 mmHg |

\# Thrombolytic Therapy Consideration

Based on the information provided, the patient may be a candidate for thrombolytic therapy (such as tPA) given the following considerations:

- Time Window: The patient presented within the 4.5-hour window for thrombolytic therapy.

- CT Findings: No hemorrhage or significant hypodensity, which supports the use of thrombolytics.

- Blood Pressure: Although elevated, blood pressure can be managed to meet the criteria for thrombolytic therapy.

- Medical History: No contraindications such as recent major surgery, major bleeding, or cancer.

\# Recommendations

- Blood Pressure Management: Before administering thrombolytics, the patient's blood pressure should be lowered to below 185/110 mmHg, as per guidelines.

- Thrombolytic Therapy: If blood pressure is controlled, and no other contraindications are present, thrombolytic therapy can be considered.

It is crucial to follow hospital protocols and consult with a stroke specialist when making the final decision.

2. OpenAI 4o (Structured Prompting) - Case A

\# Extract Critical Information

| Category | Details |
| --- | --- |
| Patient Demographics | 55-year-old woman |
| Symptom Onset | 75 minutes prior to ED arrival |
| Current Time of Evaluation | 75 minutes post-onset |
| Neurological Deficits | Right facial droop, right arm weakness, NIHSS score of 9 |
| Vital Signs | Blood pressure: 190/115 mmHg |
| Lab Values | Blood glucose: 125 mg/dL |
| Imaging Findings | CT head: No hemorrhage or frank hypodensity |
| Medical History | Well-controlled hypertension |
| Medications | Not specified |

\# Time Analysis

| Factor | Details |
| --- | --- |
| Time Since Symptom Onset | 75 minutes |
| Applicable Time Window | 0-3 hours |

\# Contraindication Check

| Type | Details | Class of Recommendation | Level of Evidence |
| --- | --- | --- | --- |
| Absolute Contraindications | None identified | N/A | N/A |
| Relative Contraindications | Elevated blood pressure (190/115 mmHg) | Class I | Level of Evidence B |

\# Decision Process

- Recommendation: Yes, recommend thrombolysis.

- Reasoning: The patient is within the 0-3 hour window for thrombolytic therapy, has no absolute contraindications, and the elevated blood pressure can be managed with antihypertensive agents to meet the criteria for safe administration of alteplase (Class I; Level of Evidence B).

\# Risk/Benefit Discussion

"Based on the current guidelines, administering thrombolytic therapy (alteplase) is recommended for you. This treatment can significantly improve your chances of recovery by dissolving the clot causing your stroke. However, it does come with a risk of bleeding, including in the brain, which can be serious. Your blood pressure is currently high, but we can manage it with medication to safely proceed with the treatment. The benefits of receiving alteplase within the first 3 hours of stroke onset generally outweigh the risks, especially given your current symptoms and the absence of major contraindications. We will closely monitor your condition and adjust treatment as necessary."

3. OpenAI o3 (Simple Prompting) - Case A

####Is Tina Patel a Candidate for IV Thrombolysis?

Yes—provided her blood pressure is lowered to ≤185/≤110 mm Hg, she meets American Heart Association/American Stroke Association (AHA/ASA) criteria for intravenous alteplase (tPA) within 3 hours of symptom onset.

####Key Eligibility Checklist

| Criterion | AHA/ASA Threshold | Patient’s Data | Meets Criterion? |
| --- | --- | --- | --- |
| Time from symptom onset | ≤3 h (strong); 3–4.5 h (extended) | 75 min (1.25 h) | Yes |
| Age | ≥18 y | 55 y | Yes |
| Neurologic deficit | Measurable | NIHSS 9 | Yes |
| Head CT | No hemorrhage/major infarct | No hemorrhage, no frank hypodensity | Yes |
| Glucose | 50–400 mg/dL | 125 mg/dL | Yes |
| Active internal bleeding, coagulopathy, recent surgery, cancer | None present | None | Yes |
| Blood pressure before tPA | ≤185 systolic AND ≤110 diastolic | 190/115 mm Hg | No (needs treatment) |

####Management Steps

1. Rapid BP lowering

• IV labetalol 10–20 mg over 1–2 min (may repeat once) OR

• Start IV nicardipine 5 mg/h, titrate by 2.5 mg/h q5 min (max 15 mg/h)

Goal: <185/<110 mm Hg prior to bolus and maintain <180/<105 mm Hg during/after infusion.

2. Re-check BP; if target achieved and no new contraindication arises, proceed with alteplase 0.9 mg/kg (max 90 mg; 10 % bolus over 1 min, remainder over 60 min).

3. Post-tPA care

• Neuro and BP checks q15 min ×2 h, q30 min ×6 h, then q1 h ×16 h.

• No antiplatelet/anticoagulant for 24 h; repeat CT/MR at 24 h before starting.

####Why Thrombolysis Is Appropriate

• Time window: She is well within the 3-hour high-benefit window.

• Moderate stroke severity (NIHSS 9) with disabling deficits (face/arm weakness).

• No absolute exclusion criteria.

• Hypertension is treatable; once controlled, BP is no longer a contraindication.

####Bottom Line

Offer IV alteplase after promptly lowering her blood pressure below 185/110 mm Hg. The potential functional benefit outweighs the bleeding risk in this scenario.

4. OpenAI o3 (Structured Prompting) - Case A

\# Extracted Critical Information

| Category | Key Findings | Relevance to tPA Decision |
| --- | --- | --- |
| Patient | 55-year-old woman | Age ≥18 → eligible (Guideline: Age, Class I, LOE A) |
| Symptom onset | Sudden right facial droop & arm weakness; onset documented 75 min before ED arrival | Within 0-3 h window; strongest benefit (Class I, LOE A) |
| Current evaluation time | Immediately on ED arrival (door-to-needle not yet achieved) | Still <3 h since onset |
| NIHSS | 9 (moderate, disabling) | Meets “treat disabling stroke” criterion (Class I, LOE A) |
| CT head | No hemorrhage, no frank hypodensity | Meets imaging safety requirement (Class I, LOE C; Class I, LOE A re: no large hypoattenuation) |
| Blood glucose | 125 mg/dL | Within safe range (Class I, LOE A) |
| Blood pressure | 190/115 mmHg | Above 185/110 threshold; may be lowered (Guideline: Control Hypertension, Class I, LOE B) |
| Platelets / INR / aPTT | Not provided; no history suggesting coagulopathy | Reasonable to proceed without delay while labs pending (Class IIa, LOE B) |
| Med‐surg history | Well-controlled HTN; no surgery, bleeding, cancer, or anticoagulants | No contraindications identified |
| Medications | None anticoagulant/antiplatelet listed | No contraindication |
| Pregnancy/others | Not pregnant (implied) | — |

\# Time Analysis

• Symptom onset → now: 75 min (1 h 15 min).

• Applicable window: 0–3 h (best-evidence window).

\# Contraindication Checklist

Absolute contraindications assessed:

– Intracranial hemorrhage? No (CT negative).

– BP uncontrolled >185/110? Currently yes, but controllable → not absolute if treatable (Class I, LOE B).

– Coagulopathy, platelets <100 k, INR > 1.7, aPTT > 40? No data suggesting; low suspicion.

– Recent head trauma, intracranial surgery, MI, GI bleed, etc.? None reported.

No absolute contraindication found.

Relative considerations: need to lower BP first and confirm stability <185/110.

\# Decision Process

Meets all eligibility criteria once BP lowered. Benefits of treatment at 75 min are high, and no contraindications exist.

\# Recommendation

Yes – Offer intravenous alteplase (tPA) after safely lowering and stabilizing BP to <185/110 mmHg, then maintain <180/105 mmHg for 24 h.

\# Supporting Guideline Citations

• Treat within 3 h if eligible (Class I; LOE A).

• Moderate or disabling stroke symptoms warrant treatment (Class I; LOE A).

• BP may be safely lowered to <185/110 before alteplase (Class I; LOE B).

• Age ≥18 yrs: no upper-age exclusion (Class I; LOE A).

• Negative CT for hemorrhage is required (Class III harm if hemorrhage).

If BP could not be lowered/stabilized, alteplase would be contraindicated; in that scenario proceed with antiplatelet therapy and consider mechanical thrombectomy if large-vessel occlusion is found.

\# Risk / Benefit Discussion (provider-to-patient speech)

“Mrs. Patel, you are having a moderate stroke that is affecting the right side of your face and arm. We are still early—about an hour and fifteen minutes since symptoms began—so you are within the window where the clot-dissolving medicine called alteplase (tPA) has the greatest chance of helping. In people treated this early, about 1 out of 3 achieves a much better recovery, sometimes returning close to normal function.

The main risk is bleeding inside the brain; this happens in about 2–3 % of treated patients and can occasionally be serious. Your current blood pressure is a little high, so we will give medicine to bring it below 185/110 first, which lowers the bleeding risk. Your scan shows no bleeding, your blood sugar is normal, and you have no other medical issues that increase the danger.

Overall, we believe the potential benefit of reducing or reversing your disability clearly outweighs the bleeding risk. If you agree, we can start the blood-pressure medication right away and give the clot-buster as soon as the pressure is safely controlled, keeping everything on schedule to give you the best chance at recovery. Do you have any questions or concerns before we proceed?”

5. Meta Llama-4-Scout-17B-16E-Instruct (Simple Prompting) - Case A

To determine if Tina Patel should be offered thrombolytics, let's consider the key points from her presentation and the guidelines for thrombolytic therapy in acute ischemic stroke:

1. Time from Symptom Onset: Tina presented 75 minutes after the onset of symptoms. Thrombolytic therapy with intravenous tissue plasminogen activator (tPA) is most effective when administered within 60 minutes of symptom onset but can be considered up to 4.5 hours after symptom onset in selected patients.
2. NIH Stroke Scale (NIHSS) Score: Her NIHSS score is 9, indicating a moderate stroke. The NIHSS score helps in assessing the severity of the stroke but does not alone determine eligibility for thrombolytics.
3. Blood Glucose: Her blood glucose is 125 mg/dL, which is slightly elevated but not in a range that typically contraindicates thrombolytic therapy.
4. CT Head Findings: The CT head showed no hemorrhage or frank hypodensity. The absence of hemorrhage is crucial because thrombolytics are contraindicated in hemorrhagic strokes.
5. Blood Pressure: Her blood pressure is currently 190/115 mmHg. Elevated blood pressure is common in acute stroke. According to guidelines, thrombolytic therapy can be administered if the blood pressure is ≤185/110 mmHg.
6. History of Major Surgery, Major Bleeding, or Cancer: She has no history of these conditions, which are important to consider due to their association with an increased risk of bleeding.

Given these points:

- Time: She is within the therapeutic time window for thrombolytic therapy (75 minutes is within the 4.5-hour window and well within the optimal early time frame).
- CT Findings: No contraindications (e.g., hemorrhage) are seen on the CT.
- Blood Pressure: It is slightly elevated but needs to be managed before administering thrombolytics. According to AHA guidelines, if the blood pressure exceeds 185/110 mmHg, thrombolytics should not be administered until it is lowered.

Management:

Before offering thrombolytics, her blood pressure needs to be carefully managed and lowered to ≤185/110 mmHg. Once her blood pressure is within the acceptable range and after re-evaluation for any changes in her neurological status or new contraindications, thrombolytic therapy could be considered. Close monitoring of her blood pressure and neurological status is crucial during and after the administration of thrombolytics.

In conclusion, provided that her blood pressure can be safely lowered to ≤185/110 mmHg, Tina Patel could be offered thrombolytics given her eligibility based on time from symptom onset, NIHSS score, and absence of hemorrhage on CT head. However, her elevated blood pressure requires careful management before the administration of thrombolytic therapy.

6. Llama-4-Scout-17B-16E-Instruct (Structured Prompting) - Case A

**Step 1: Extract Critical Information**

- Age: 55 years old
- Time of symptom onset: Not specified, but patient presented 75 minutes after onset
- Current time of evaluation: Not specified, but 75 minutes after symptom onset
- Neurological deficits: Right facial droop and right arm weakness, NIHSS score of 9
- Vital signs: Blood pressure 190/115 mmHg
- Lab values: Glucose 125 mg/dL, no mention of platelets, INR, or other lab values
- Imaging findings: CT head showed no hemorrhage or frank hypodensity
- Medical history: Well-controlled hypertension, no history of major surgery, major bleeding, or cancer

**2: Time Analysis**

- Time since symptom onset: 75 minutes (1 hour 15 minutes)
- The patient is within the 3-hour window for thrombolytic therapy consideration.

**3: Contraindication Check**

- Absolute contraindications:
  - No mention of active bleeding, recent surgery, or history of intracranial hemorrhage.
  - Blood pressure is elevated at 190/115 mmHg.
- Relative contraindications:
  - Elevated blood pressure (190/115 mmHg) which needs to be controlled before administering tPA.

**4: Decision Process**

- The patient is within the 3-hour window.
- No absolute contraindications mentioned except for the need to control blood pressure.
- Elevated blood pressure is a concern but can be managed.

**5: Guidelines Application**

- According to guidelines, intravenous alteplase is recommended for patients whose blood pressure can be lowered safely to <185/110 mmHg with antihypertensive agents before starting treatment (Class I; Level of Evidence B).

**6: Recommendation**

Given the patient's presentation and the need to control her blood pressure, the recommendation would be to attempt to lower her blood pressure to <185/110 mmHg and then consider thrombolytic therapy.

**7: Risk/Benefit Discussion**

The patient and family should be informed about the risks and benefits of thrombolytic therapy, including the risk of symptomatic intracranial hemorrhage and the potential for significant improvement in neurological deficits.

**8: Final Decision**

The patient should be offered thrombolytics after blood pressure control.

The best answer is:

Yes

7. Llama-3.3-70B-Instruct-Turbo (Simple Prompting) - Case A

To determine if Tina Patel should be offered thrombolytics, we need to consider the guidelines for thrombolytic therapy in acute ischemic stroke. The American Heart Association (AHA) and American Stroke Association (ASA) recommend thrombolytic therapy with intravenous recombinant tissue plasminogen activator (tPA) for eligible patients with acute ischemic stroke.

The key criteria for thrombolytic therapy include:

1. Time of onset: The patient presented within 4.5 hours of symptom onset (in this case, 75 minutes).
2. Neurological deficit: The patient has a significant neurological deficit, as indicated by an NIHSS score of 9.
3. Imaging: The CT head shows no hemorrhage or frank hypodensity, making it safe to administer thrombolytics.
4. Blood pressure: The patient's blood pressure is elevated (190/115 mmHg), but this is not an absolute contraindication. The AHA/ASA guidelines suggest that blood pressure should be below 185/110 mmHg before administering tPA. However, if the blood pressure can be safely lowered to this range, thrombolytics can still be considered.
5. Other contraindications: The patient has no history of major surgery, major bleeding, or cancer, which are other potential contraindications to thrombolytic therapy.

Given these factors, Tina Patel is a candidate for thrombolytic therapy. However, her elevated blood pressure needs to be addressed before administering tPA. The emergency department team should consider lowering her blood pressure to a safe range (<185/110 mmHg) using antihypertensive medications, such as labetalol or nicardipine, before offering thrombolytics.

In summary, Tina Patel should be considered for thrombolytic therapy after her blood pressure is safely lowered to an acceptable range. Close monitoring and management of her blood pressure and other vital signs are crucial during the thrombolytic therapy.

8. Llama-4-Scout-17B-16E-Instruct (Structured Prompting) - Case A

To determine whether thrombolytic therapy (tPA/alteplase) should be offered to Tina Patel, we need to systematically analyze her case based on the provided guidelines.

**Extract Critical Information**

1. Patient Demographics: Tina Patel is a 55-year-old woman.
2. Precise Time of Symptom Onset: Symptoms started 75 minutes before presentation to the emergency department.
3. Current Time of Evaluation: Not explicitly stated, but given the symptom onset, it's within the 3-hour window if evaluated immediately upon arrival.
4. Neurological Deficits: Right facial droop and right arm weakness, with an NIHSS score of 9.
5. Vital Signs: Blood pressure is 190/115 mmHg.
6. Relevant Lab Values: Blood glucose is 125 mg/dL.
7. Imaging Findings: CT head showed no hemorrhage or frank hypodensity.
8. Medical History and Medications: Well-controlled hypertension, no history of major surgery, major bleeding, or cancer.

**Time Analysis**

1. Exact Time Since Symptom Onset: 75 minutes.
2. Time Window: Within the 0-3 hour window, which is the primary time frame for considering thrombolytic therapy.

**Contraindication Check**

1. Absolute Contraindications: None mentioned (e.g., no intracranial hemorrhage, no recent head trauma, no intracranial neoplasm, no known coagulopathies or bleeding diatheses).
2. Relative Contraindications: Elevated blood pressure (190/115 mmHg), which can be managed.
3. Special Circumstances: None mentioned that would require additional consideration.

**Decision Process**

Given that Tina Patel is within the 3-hour window and has no absolute contraindications that cannot be quickly corrected (her elevated blood pressure can be managed), the recommendation would lean towards thrombolysis. However, her elevated blood pressure needs to be addressed before administering tPA, as guidelines recommend blood pressure to be below 185/110 mmHg (Class I; Level of Evidence B).

**Response Format**

Recommendation: Yes, with the condition that her blood pressure is safely lowered to the recommended levels before administering tPA.

Supporting Reasoning: The patient's presentation within the 3-hour window, lack of absolute contraindications, and the potential to manage her elevated blood pressure align with guidelines supporting the use of thrombolytic therapy for acute ischemic stroke (Class I; Level of Evidence A).

Risk/Benefit Discussion Speech:

"Tina, based on your symptoms and our evaluation, you are a candidate for a treatment called thrombolytic therapy, which can help dissolve the clot causing your stroke. However, we need to manage your high blood pressure first to ensure it's safe to proceed. The benefits of this treatment include improving outcomes and reducing the risk of long-term disability from your stroke. There are risks, including a small chance of bleeding in the brain, but we believe the benefits outweigh these risks in your case. We will closely monitor you and adjust your treatment as necessary. Do you have any questions or concerns about this treatment?"

9. OpenAI 4o (Simple Prompting) - Case B

\# Patient Information

| Parameter | Details |
| --- | --- |
| Age | 78 years old |
| Medical History | Atrial fibrillation (on warfarin), Type 2 diabetes |
| Current Symptoms | Aphasia, severe left-sided weakness |
| Time Last Seen Normal | 3 hours ago |
| NIHSS Score | 28 |
| INR | 1.8 |
| Blood Pressure | 175/90 mmHg |
| CT Head Findings | Early subtle ischemic changes, no significant hypodensity or hemorrhage |
| Recent Medical History | Basal cell carcinoma excised last week |

\# Thrombolytic Therapy Consideration

| Factor | Consideration |
| --- | --- |
| Time Window | Within 4.5 hours for thrombolytic therapy eligibility |
| INR Level | 1.8, which is below the typical threshold of 1.7 for safe thrombolytic use in patients on warfarin |
| Blood Pressure | 175/90 mmHg, which may need management but is not an absolute contraindication |
| CT Findings | No hemorrhage, which supports eligibility |
| Recent Surgery | Basal cell carcinoma excision last week, generally not considered major surgery |

\# Conclusion

Based on the provided information, the patient may be considered for thrombolytic therapy, as he is within the 4.5-hour window, has an INR of 1.8, and no contraindications such as significant hemorrhage or recent major surgery. However, the decision should be made in consultation with a stroke specialist, considering all clinical factors and potential risks.

10. OpenAI 4o (Structured Prompting) - Case B

\# Extract Critical Information

| Category | Details |
| --- | --- |
| Patient Demographics | 78-year-old male |
| Time of Symptom Onset | Last seen normal 3 hours ago |
| Current Time of Evaluation | Within 3-hour window |
| Neurological Deficits | Aphasia, severe left-sided weakness, NIHSS score of 28 |
| Vital Signs | Blood pressure: 175/90 mmHg |
| Lab Values | INR: 1.8 |
| Imaging Findings | CT head shows early subtle ischemic changes, no significant hypodensity or hemorrhage |
| Medical History and Medications | Atrial fibrillation (on warfarin), type 2 diabetes, recent basal cell carcinoma excision |

\# Time Analysis

- Time since symptom onset: 3 hours

- Applicable time window: 0-3 hours

\# Contraindication Check

- Absolute Contraindications:

- INR >1.7 is a contraindication for thrombolysis (Class III; Level of Evidence C).

- Relative Contraindications:

- Recent surgery (basal cell carcinoma excision last week) is not considered major surgery and does not pose a significant risk.

- Elevated blood pressure can be managed to meet criteria for thrombolysis (Class I; Level of Evidence B).

\# Decision Process

- Recommendation: No

- Reasoning: The patient's INR is 1.8, which exceeds the recommended threshold of ≤1.7 for safe administration of thrombolytics. This is an absolute contraindication according to the guidelines (Class III; Level of Evidence C).

\# Alternative Management

- Consider mechanical thrombectomy if applicable and within the appropriate time window.

- Initiate supportive care and secondary prevention measures, including blood pressure management and anticoagulation adjustment.

\# Risk/Benefit Discussion Speech

"Mr. Mercado, based on the current guidelines and your medical condition, administering thrombolytic therapy is not recommended due to your INR level being above the safe threshold. This increases the risk of bleeding complications. However, we will focus on other treatment options, such as managing your blood pressure and considering mechanical thrombectomy if suitable. Our goal is to minimize the impact of the stroke and support your recovery. We will continue to monitor your condition closely and adjust your treatment plan as needed."

11. OpenAI o3 (Simple Prompting) - Case B

\# Assessment of IV Thrombolysis (Alteplase/Tenecteplase) Eligibility

| Factor | Patient Data | Guideline Threshold/Comment | Does it Exclude tPA? |
| --- | --- | --- | --- |
| Time from last known well | ≈3 hours | ≤4.5 hours (3 h class I; 3–4.5 h class II) | No |
| Neurologic deficit severity | NIHSS 28 | Severity itself does not preclude tPA | No |
| Blood pressure | 175/90 mmHg | Must be <185/110 mmHg before bolus | No (already below) |
| Anticoagulation (warfarin) | INR 1.8 | Absolute exclusion if INR > 1.7 or PT > 15 s | YES |
| Recent surgery/trauma | Minor skin excision 7 days ago | Minor surgery in last 14 days – relative risk, not absolute | No (but note caution) |
| Imaging | Early subtle changes, no hemorrhage | No frank hypodensity/hemorrhage → acceptable | No |

\# Guideline Synthesis

• 2018 AHA/ASA acute ischemic stroke guidelines list “treatment with warfarin and INR > 1.7” as an absolute contraindication (Class III: Harm).

• Minor skin procedures within 14 days are only relative contraindications, but they become moot once an absolute contraindication is present.

• Blood pressure is within the treatable range, and imaging is permissible.

\# Recommendation

Because the INR is 1.8 (>1.7), Joseph Mercado does NOT meet safety criteria for IV thrombolytic therapy. He should therefore not receive alteplase or tenecteplase.

\# Next Steps

• Urgently obtain vascular imaging (CTA/MRA) to determine candidacy for mechanical thrombectomy (especially given NIHSS 28 suggesting large-vessel occlusion).

• Continue standard stroke care: maintain BP <180/105 mmHg, reverse anticoagulation only if needed for thrombectomy, manage glucose, monitor airway/swallowing, and admit to a stroke unit or ICU.

12. OpenAI o3 (Structured Prompting) - Case B

\# Extracted Critical Information

| Category | Details | Notes |
| --- | --- | --- |
| Patient demographics | 78-year-old man | Age ≥18 (eligible age group) |
| Symptom onset | Last known normal 3 h ago | Precise time given |
| Current evaluation time | Within 3 h window | Falls inside 0-to-3-h window |
| Neurologic deficits | Global aphasia, dense left-sided weakness, NIHSS 28 | Severe, clearly disabling |
| Vital signs | BP 175/90 mmHg | Below 185/110; can treat if needed |
| Laboratory values | INR 1.8; platelets, glucose not stated | INR >1.7 (key) |
| Imaging | Non-contrast CT: early subtle ischemic change, no hemorrhage, no large frank hypodensity | Meets imaging safety criteria |
| History/medications | Chronic atrial fibrillation on warfarin; type 2 diabetes; basal-cell carcinoma excised 7 days ago | Anticoagulation is the critical issue |

\# Time Analysis

• Onset-to-door: ≈3 h → within standard 0-to-3-h treatment window.

\# Contraindication Check

| Potential Contraindication | Present? | Guideline reference & strength |
| --- | --- | --- |
| INR > 1.7 while on warfarin | Yes (1.8) | Absolute: “INR > 1.7 … IV alteplase is not recommended” (Class III; LOE B) |
| Recent surgery <14 days | Minor skin excision 7 days ago | Relative: “Major surgery” within 14 d (Class IIb; LOE C). Minor dermatologic excision ≠ major, but moot given INR. |
| BP >185/110 | No | Can be controlled (Class I; LOE B). |
| Other absolute exclusions (ICH, platelets <100k, glucose <50 or >400, etc.) | No data suggesting | Not present. |

Absolute contraindication confirmed: warfarin with INR 1.8 (>1.7).

\# Decision Process

Because an absolute contraindication is present, intravenous alteplase should NOT be administered.

Recommendation: No.

Class III; Level of Evidence B—administration would be potentially harmful.

\# Alternative / Adjunctive Management

• Obtain CT/CTA to assess for large-vessel occlusion; if present and within 0–24 h criteria, pursue mechanical thrombectomy.

• Initiate aggressive secondary-prevention measures: resume anticoagulation when safe, control glucose and BP.

• Begin antiplatelet therapy (e.g., aspirin) 24 h after ruling out hemorrhage if anticoagulation must be delayed.

• Admit to stroke unit; monitor for neurological or hemorrhagic deterioration.

\# Provider-to-Patient (or Family) Risk/Benefit Discussion

“Mr. Mercado is having a severe stroke. One therapy we often use is the clot-busting drug alteplase (sometimes called tPA). This medicine works best when given within 3 hours, but it carries a risk of dangerous bleeding in the brain.

Because you take warfarin and your blood test today shows your clotting level (INR) is 1.8, current national stroke guidelines tell us that giving alteplase would place you at an unacceptably high risk of bleeding with little proven benefit. Therefore, we do not recommend giving the drug.

What we will do instead is focus immediately on other treatments. We will get additional scans to see if a catheter-based retrieval procedure is possible, tightly control your blood pressure and blood sugar, and admit you to our specialized stroke unit for close monitoring and supportive care. We will also plan the safest timing to restart your anticoagulation for your atrial fibrillation.

If you have any questions about these recommendations or alternatives, please let us know—we want to make sure you feel fully informed and comfortable with the plan.”

13. Meta Llama-4-Scout-17B-16E-Instruct (Simple Prompting) - Case B

**Step 1: Assess the patient's eligibility for thrombolytics based on the time window**

The patient was last seen normal 3 hours ago and presents with symptoms of a stroke, making him within the potential time window for thrombolytic therapy, which is typically considered within 4.5 hours of symptom onset for eligible patients.

**2: Evaluate the patient's NIHSS score**

The patient's NIHSS (National Institutes of Health Stroke Scale) score is 28, indicating severe stroke symptoms. A high NIHSS score suggests significant neurological deficit but does not directly contraindicate thrombolysis.

**3: Consider the patient's INR level**

The patient's current INR (International Normalized Ratio) is 1.8. For patients on warfarin, an INR ≤ 1.7 is often considered acceptable for safe administration of alteplase (a thrombolytic agent), though guidelines may slightly vary.

**4: Assess blood pressure eligibility**

The patient's blood pressure is 175/90 mmHg. Generally, thrombolytics can be administered if the blood pressure is ≤ 185/110 mmHg. Therefore, this patient's blood pressure is slightly elevated but within a range that requires close monitoring rather than an absolute contraindication.

**5: Evaluate the CT head findings**

The CT head shows early subtle ischemic changes but no significant hypodensity or hemorrhage. The absence of significant hypodensity or hemorrhage on CT is crucial for thrombolytic eligibility, as it suggests the ischemic changes are early and not yet established.

**6: Consider recent surgery and bleeding history**

The patient had basal cell carcinoma excised last week. Recent surgery, especially major surgery, can be a contraindication to thrombolytics, but the excision of basal cell carcinoma is generally considered minor and might not pose a significant risk.

**7: Assess the presence of atrial fibrillation and warfarin use**

The patient is on warfarin for atrial fibrillation. Despite the slightly subtherapeutic INR of 1.8, the primary concern with thrombolytics in this context is not the indication for anticoagulation but ensuring there's no contraindication to its use.

**8: Determine eligibility based on guidelines**

Given the patient's presentation within 3 hours of symptom onset, severe stroke symptoms (NIHSS 28), INR of 1.8, blood pressure of 175/90 mmHg, and absence of significant CT abnormalities or recent major bleeding, the patient appears to be a candidate for thrombolytic therapy. The recent minor surgery (basal cell carcinoma excision) and slightly elevated blood pressure are noted but do not clearly contraindicate treatment.

The best answer is Yes.

14. Meta Llama-4-Scout-17B-16E-Instruct (Structured Prompting) - Case B

**Step 1: Extract Critical Information**

- Patient demographics: 78-year-old man
- Time of symptom onset: 3 hours ago
- Current time of evaluation: Not specified, but implied to be within 3 hours of symptom onset
- Neurological deficits: Aphasia and severe left-sided weakness, NIHSS score 28
- Vital signs: Blood pressure 175/90 mmHg
- Lab values: INR 1.8, platelets not specified, glucose not specified
- Imaging findings: CT head shows early subtle ischemic changes but no significant hypodensity or hemorrhage
- Medical history: Atrial fibrillation (on warfarin), type 2 diabetes
- Recent history: Basal cell carcinoma excised last week, no recent major surgery or major bleeding

**2: Time Analysis**

- Time since symptom onset: 3 hours
- Time window: Within 3 hours of symptom onset, which is a recommended time frame for thrombolytic therapy.

**3: Contraindication Check**

- Absolute contraindications:
  - No clear evidence of active bleeding or severe coagulopathy (INR 1.8 is slightly elevated but not drastically high)
  - No recent major surgery or major bleeding
  - No history of intracranial hemorrhage
  - No severe head trauma within 3 months
  - No known coagulopathies or bleeding diatheses that would contraindicate tPA
- Relative contraindications:
  - Elevated blood pressure (175/90 mmHg), but it can be managed
  - Recent basal cell carcinoma excision (last week), which might pose a slight risk but is not a strong contraindication
  - Age (78 years) is not a contraindication but a factor to consider in risk-benefit analysis

**4: Decision Process**

- The patient is within the 3-hour window, which is favorable for thrombolytic therapy.
- There are no clear absolute contraindications listed.
- The patient's INR is 1.8, which is slightly elevated but not a strong contraindication for tPA administration in this context.
- The patient's severe stroke symptoms (NIHSS 28) suggest a significant potential benefit from thrombolytic therapy.

**5: Recommendation**

Given the patient's severe stroke symptoms, the time since symptom onset, and the absence of clear absolute contraindications, thrombolytic therapy with tPA is recommended.

**6: Risk/Benefit Discussion**

"The benefits of thrombolytic therapy in your case include a potential significant improvement in your neurological deficits and reduction in long-term disability. However, there are risks, including a small chance of symptomatic intracranial hemorrhage. Given your severe stroke symptoms and the fact that you are within the 3-hour treatment window, we believe the potential benefits outweigh the risks. We will closely monitor you for any signs of complications."

The best answer is Yes.

15. Llama-3.3-70B-Instruct-Turbo (Simple Prompting) - Case B

To determine if this patient should be offered thrombolytics, we need to consider the indications and contraindications for thrombolytic therapy in acute ischemic stroke.

Indications for thrombolytic therapy with intravenous recombinant tissue plasminogen activator (tPA) include:

1. Diagnosis of acute ischemic stroke
2. Time of onset of symptoms ≤ 4.5 hours (or time last seen normal)
3. NIHSS score ≥ 4 (indicating significant stroke severity)

This patient meets these criteria, as he has a diagnosis of acute ischemic stroke (suggested by aphasia and severe left-sided weakness), the time of onset is ≤ 4.5 hours (last seen normal 3 hours ago), and his NIHSS score is 28, indicating severe stroke.

Contraindications for thrombolytic therapy include:

1. Significant head trauma or stroke in the preceding 3 months
2. Intracranial hemorrhage or suspicion of hemorrhage
3. Recent major surgery (within 14 days)
4. Recent gastrointestinal or urinary tract hemorrhage (within 21 days)
5. Severe uncontrolled hypertension (typically systolic > 185 mmHg or diastolic > 110 mmHg)
6. Elevated INR > 1.7 (indicating anticoagulation)
7. Platelet count < 100,000/μL

This patient has an INR of 1.8, which is slightly elevated, indicating that he is anticoagulated on warfarin. However, the INR is not significantly elevated, and the patient's current clinical situation may outweigh the risks associated with a mildly elevated INR.

The patient's blood pressure is 175/90 mmHg, which is elevated but not significantly above the typical threshold for withholding thrombolytics.

The recent excision of basal cell carcinoma is not a contraindication for thrombolytic therapy, as it is not considered a major surgery.

Considering the patient's clinical presentation, the benefits of thrombolytic therapy likely outweigh the risks. Therefore, this patient should be considered for thrombolytic therapy, and the decision should be made in consultation with a neurologist or stroke specialist. It is essential to carefully weigh the potential benefits and risks, including the possibility of bleeding complications due to the patient's anticoagulation status.

16. Llama-3.3-70B-Instruct-Turbo (Structured Prompting) - Case B

**Extract Critical Information**

1. Patient Demographics: Joseph Mercado, a 78-year-old man.
2. Precise Time of Symptom Onset: 3 hours ago.
3. Current Time of Evaluation: Not specified, but implied to be shortly after the 3-hour mark since symptom onset.
4. Neurological Deficits: Aphasia and severe left-sided weakness, with an NIHSS score of 28.
5. Vital Signs: Blood pressure is 175/90 mmHg.
6. Relevant Lab Values: INR is 1.8, indicating the patient is on warfarin but within a potentially acceptable range for thrombolysis.
7. Imaging Findings: CT head shows early subtle ischemic changes but no significant hypodensity or hemorrhage.
8. Medical History and Medications: Atrial fibrillation (on warfarin), type 2 diabetes, and recent basal cell carcinoma excision.

**Time Analysis**

1. Exact Time Since Symptom Onset: 3 hours.
2. Time Window: Within the 0-3 hour window, which is the primary time frame for considering thrombolytic therapy.

**Contraindication Check**

1. Absolute Contraindications:
   - None explicitly mentioned that cannot be quickly corrected. The patient's INR of 1.8 is below the threshold of 1.7, which is often considered for thrombolysis in patients on warfarin.
   - Recent surgery (basal cell carcinoma excision) might be a concern, but guidelines suggest considering thrombolysis on a case-by-case basis for patients who have undergone major surgery in the preceding 14 days.
2. Relative Contraindications:
   - Age (78 years old) is not a contraindication according to guidelines, which state that older age is an adverse prognostic factor but does not modify the treatment effect of thrombolysis (Class I; Level of Evidence A).
   - Blood pressure is elevated but can be managed according to guidelines (Class I; Level of Evidence B).
   - The patient is on warfarin, but with an INR of 1.8, which may be considered acceptable for thrombolysis (Class IIb; Level of Evidence B).
3. Special Circumstances:
   - Recent basal cell carcinoma excision is noted, but this is not typically a contraindication for thrombolytic therapy.

**Decision Process**

Given the information:

- The patient is within the 3-hour window, which is the primary time frame for thrombolysis.
- There are no absolute contraindications that cannot be quickly corrected.
- Relative contraindications and special circumstances are present but do not necessarily preclude thrombolysis.

**Recommendation**

Yes, thrombolytic therapy should be considered for Joseph Mercado, given his presentation within the 3-hour window, the absence of absolute contraindications, and the potential benefits outweighing the risks based on the guidelines.

**Risk/Benefit Discussion**

"Mr. Mercado, you have presented with severe stroke symptoms, including aphasia and left-sided weakness, within a time frame where thrombolytic therapy can be considered. While you are on warfarin and have an elevated blood pressure, these factors can be managed. The benefits of thrombolysis in your case, given the severity of your symptoms and the potential for significant improvement, outweigh the risks. However, it's essential to discuss the potential risks, including bleeding, which is a concern with any anticoagulant therapy. Given your recent surgery, we will monitor you closely. We believe that the potential benefits of thrombolytic therapy in improving your outcome and reducing disability from this stroke make it a reasonable option for you."

**Alternative Management**

If thrombolysis were not considered due to unforeseen contraindications or complications, alternative management would include supportive care, possible consideration of mechanical thrombectomy if eligible, and management of underlying conditions such as atrial fibrillation and diabetes to prevent future strokes.

17. OpenAI 4o (Simple Prompting) - Case C

\# Thrombolytic Therapy Consideration

| Factor | Details | Consideration for Thrombolytics |
| --- | --- | --- |
| Age | 42 years old | Eligible |
| Time since symptom onset | 2 hours | Eligible (within 4.5-hour window) |
| NIHSS Score | 6 | Eligible (indicates moderate stroke) |
| Intracranial Aneurysm | 8 mm unruptured | Caution advised, but not an absolute contraindication |
| Current Menstruation | Normal flow | Not a contraindication |
| Blood Pressure | 140/80 mmHg | Eligible (within acceptable range) |
| CT Head | Negative for hemorrhage | Eligible |
| History of Intracranial Hemorrhage | None | Eligible |
| Recent Major Surgery or Cancer | None | Eligible |

\# Conclusion

Based on the provided information, Maria Gonzales can be considered for thrombolytic therapy. However, the presence of an unruptured intracranial aneurysm requires careful consideration and consultation with a neurologist or stroke specialist to weigh the risks and benefits.

18. OpenAI 4o (Structured Prompting) - Case C

\# Extract Critical Information

| Category | Details |
| --- | --- |
| Patient Demographics | 42-year-old woman |
| Symptom Onset | 2 hours ago |
| Current Time of Evaluation | 2 hours post-onset |
| Neurological Deficits | Dysarthria, difficulty walking, left-sided incoordination, NIHSS score of 6 |
| Vital Signs | Blood pressure: 140/80 mmHg |
| Lab Values | Not provided |
| Imaging Findings | CT head negative for hemorrhage |
| Medical History | Small lacunar stroke 2 months ago, 8 mm unruptured intracranial aneurysm, currently menstruating |
| Medications | Not provided |

\# Time Analysis

| Factor | Details |
| --- | --- |
| Time Since Symptom Onset | 2 hours |
| Applicable Time Window | 0-3 hours |

\# Contraindication Check

| Type | Details | Class of Recommendation | Level of Evidence |
| --- | --- | --- | --- |
| Absolute Contraindications | None identified | N/A | N/A |
| Relative Contraindications | Menstruating with normal flow, unruptured intracranial aneurysm | Class IIa | Level C |

\# Decision Process

- Recommendation: Yes, recommend thrombolysis.

- Rationale: The patient is within the 0-3 hour window for thrombolytic therapy, has no absolute contraindications, and the presence of a small unruptured aneurysm and menstruation are not contraindications per guidelines. The benefits of treatment likely outweigh the risks.

\# Risk/Benefit Discussion

"Based on the current guidelines, administering thrombolytic therapy (alteplase) is recommended for you. You are within the critical 0-3 hour window since the onset of your stroke symptoms, which maximizes the potential benefits of the treatment. While you have a small unruptured aneurysm and are currently menstruating, these factors are not considered absolute contraindications. The treatment could increase the degree of menstrual flow, but the potential benefits of reducing stroke-related neurological deficits outweigh these risks. We will monitor you closely for any complications. It is important to act quickly to improve your chances of recovery."

19. OpenAI o3 (Simple Prompting) - Case C

##### **Clinical Question**

Should IV thrombolysis (alteplase/tenecteplase) be offered to a 42-year-old woman who presents 2 h after onset of an acute ischemic stroke, with an NIHSS 6, a negative CT for hemorrhage, an unruptured 8 mm ACom aneurysm, ongoing menstruation, and a prior small lacunar stroke 2 months ago?

##### **Guideline-Based Eligibility Checklist**

| **Contraindication** | **Absolute or Relative (per 2019-2023 AHA/ASA & ESO guidance)** | **Present in this patient?** | **Comment/Rationale for this case** |
| --- | --- | --- | --- |
| Intracranial hemorrhage on CT/MRI | Absolute | No | Non-contrast CT is normal. |
| Intracranial neoplasm, AVM, aneurysm ≥10 mm or recently ruptured | Absolute for ruptured; relative for unruptured <10 mm | Yes (8 mm unruptured) | Evidence from multiple series shows no excess risk with unruptured aneurysms <10 mm. |
| Ischemic stroke within prior 3 months | Relative (no longer absolute) | Yes (8 weeks ago, small lacunar) | Small, fully resolved lacunar infarct; imaging currently normal → acceptable to treat. |
| Intracranial hemorrhage history | Absolute | No | Prior stroke was ischemic only. |
| Systolic BP >185 mmHg or diastolic >110 mmHg | Absolute until controlled | No | 140/80 mmHg without antihypertensive therapy. |
| Platelets <100 000, INR > 1.7, aPTT prolonged, DOAC use <48 h | Absolute | Unknown but no history of anticoagulant use given | Assume normal if labs confirm. |
| Major surgery/serious trauma ≤14 d | Absolute | No | None reported. |
| Active internal bleeding | Absolute | No | Menstruation with normal flow is not a contraindication. |
| Suspected infective endocarditis, aortic dissection | Absolute | No | Not suspected. |
| Glucose <50 or >400 mg/dL | Absolute until corrected | No | Assume normal pending POC test. |

No absolute contraindication is present. The two relative issues (recent small stroke and asymptomatic aneurysm <10 mm) are explicitly permitted in up-to-date guidelines, provided imaging is reassuring and the treating team judges that benefits outweigh risks.

##### **Benefit–Risk Appraisal**

• Time window: Symptom onset to door is 2 h → within both 0–3 h and 3–4.5 h approved windows, maximizing benefit.
• Clinical severity: NIHSS 6 (moderate, disabling dysarthria, gait ataxia, incoordination). Functional disability is likely without treatment.
• Hemorrhagic risk:
– Prior lacunar infarct is small, now radiographically silent.
– Unruptured aneurysm <10 mm shows no excess sICH in pooled analyses.
– Menstruation has no proven added risk.
– BP in target range.
Overall estimated symptomatic intracranial hemorrhage risk ≈2–3 % (no higher than baseline). Estimated chance of significant functional improvement ~30 % when treated at 2 h vs ~15 % without thrombolysis.

##### **Recommendation**

Offer IV thrombolysis after standard informed consent. The expected functional benefit clearly outweighs the modest incremental bleeding risk associated with a small prior stroke and an unruptured 8 mm aneurysm. Mechanical thrombectomy is not indicated because no large-vessel occlusion is described, but vascular imaging should still be obtained per protocol.

20. OpenAI o3 (Structured Prompting) - Case C

##### **Extracted Case Information**

| **Category** | **Details** |
| --- | --- |
| Patient | Maria Gonzales, 42-year-old woman |
| Symptom onset | 2 h ago – witnessed, clear time |
| Current evaluation time | Within 3 h window (0-3 h) |
| Neurologic deficits | Dysarthria, gait difficulty, left incoordination; NIHSS = 6 (moderate, potentially disabling) |
| Vital signs | BP 140/80 mmHg (below 185/110 threshold) |
| Imaging | Non-contrast CT head: no hemorrhage, no large early ischemic change |
| Labs | Not provided; no reason to suspect coagulopathy |
| Past medical history | Small lacunar infarct (left basal ganglia) 2 months ago; fully recovered (mRS 0–1) |
| Medications | None reported; no anticoagulant/antiplatelet issues noted |
| Other conditions | 8 mm unruptured ACom aneurysm; currently menstruating, normal flow |
| Exclusions specifically not present | No ICH history, no recent major surgery or trauma, no cancer, BP controlled, glucose not low/high, no infective endocarditis, no aortic dissection |

##### **Time Analysis**

• Onset-to-needle candidate window = 2 h → inside 0–3 h window (maximal proven benefit).
• Door-to-needle goal ≤60 min still possible.

##### **Contraindication Check**

| **Item** | **Present?** | **Guideline note (Class/LOE)** | **Interpretation** |
| --- | --- | --- | --- |
| Prior ischemic stroke <3 mo | Yes (2 mo) | Potentially harmful to give alteplase (Class III; LoE B) BUT expert consensus allows in selected cases if small infarct, >1 mo, good recovery | Relative – warrants shared decision |
| Intracranial aneurysm <10 mm | Yes (8 mm) | IV alteplase reasonable (Class IIa; LoE C) | Not a contraindication |
| Active menstruation | Yes | Alteplase probably indicated; warn may ↑ flow (Class IIa; LoE C) | Not a contraindication |
| BP >185/110 | No | Must be <185/110 before bolus (Class I; LoE B) | Meets criterion |
| Hemorrhage on CT | No | Absolute contraindication if present (Class III) | Meets criterion |
| Platelets<100k, INR>1.7, aPTT>40 | Unknown but no suspicion | Not required to delay without suspicion (Class IIa; LoE B) | Assume ok |

No other absolute contraindications identified.

##### **Benefit–Risk Synthesis**

Benefit
• Within golden 3-hour window – Class I; LoE A for disabling or moderate strokes.
• NIHSS 6: considered disabling by many clinicians – Class I; LoE A.

Risks
• Prior stroke 2 months ago carries uncertain ↑ sICH risk (Class III; LoE B).
• Small unruptured aneurysm and normal menstruation add minor bleeding risk but both are considered acceptable (Class IIa recommendations).

Overall, guideline language implies treatment “may be harmful” yet expert consensus supports alteplase in carefully selected patients exactly like this one (small prior infarct >1 month and full recovery). Therefore, this is a relative contraindication, not absolute.

##### **Recommendation**

Requires shared decision making with modified risk/benefit discussion.

##### **Suggested Clinician Speech for Shared Decision Making**

“Mrs. Gonzales, you are having a new stroke that began about two hours ago. A medicine called alteplase can dissolve the clot and improve the chance of a good recovery if given now. In people treated this early, about 1 in 3 gain significant extra independence.

You had a very small stroke two months ago. Because it was small, you fully recovered, and more than a month has passed, national experts agree alteplase can still be offered, but the prior stroke may very slightly increase the risk of bleeding in the brain. In the average patient the serious bleeding risk is about 2-3 %; with your recent stroke it may be a little higher, though we cannot say exactly how much.

You also have a small 8 mm aneurysm and are on your period. Both are considered safe under our guidelines, but your menstrual flow might become heavier for a day or two.

Overall, acting quickly gives you the best chance to avoid long-term problems with speech and coordination. If bleeding were to occur, it could worsen your outcome and in rare cases be life-threatening. We believe the likely benefits outweigh the risks, but because of the prior stroke we want to be sure you are comfortable proceeding. Do you have any questions, and would you like to go ahead with the treatment?”

21. Meta Llama-4-Scout-17B-16E-Instruct (Simple Prompting) - Case C

**Step 1: Assess the patient's eligibility for thrombolytics based on her medical history and current condition.**

The patient, Maria Gonzales, presents with symptoms suggestive of an acute ischemic stroke, as indicated by the onset of dysarthria, difficulty walking, and left-sided incoordination, with a National Institutes of Health Stroke Scale (NIHSS) score of 6. Her symptoms started 2 hours ago, which is within the typical time window for thrombolytic therapy.

**2: Consider the patient's history of recent stroke.**

Maria had a small lacunar stroke of the left basal ganglia 2 months ago with full recovery. Generally, patients who have had a stroke within the past 3 months are at increased risk of hemorrhage if given thrombolytics, but the decision can depend on the severity and type of the previous stroke.

**3: Evaluate the presence of the unruptured intracranial aneurysm.**

The patient has an 8 mm unruptured intracranial aneurysm of the anterior communicating artery. The presence of an unruptured aneurysm may increase the risk of hemorrhage with thrombolytic therapy, but the decision to treat is often made on a case-by-case basis considering the size of the aneurysm and the risk-benefit ratio.

**4: Assess the patient's current condition and other eligibility criteria for thrombolytics.**

Her current blood pressure is 140/80 mmHg, which is within relatively safe limits for thrombolytic therapy (typically, systolic blood pressure <185 mmHg and diastolic <110 mmHg). The CT head is negative for hemorrhage, which is a critical prerequisite for administering thrombolytics.

**5: Consider the patient's menstruating status and its implications.**

The patient is currently menstruating with a normal flow. While active menstruation might be a concern for potential bleeding risks, it is not typically an absolute contraindication for thrombolytic therapy in the context of acute ischemic stroke.

**6: Weigh the risks and benefits of thrombolytic therapy in this patient.**

Given the patient's presentation within the therapeutic time window, the benefits of thrombolytic therapy in reducing disability from ischemic stroke must be weighed against the risks, including the potential for hemorrhage and the presence of an unruptured aneurysm.

**7: Determine the appropriateness of thrombolytic therapy.**

Considering the patient's recent stroke, unruptured aneurysm, and current condition, the decision to offer thrombolytics requires careful consideration. However, guidelines typically suggest that recent stroke within 3 months is a relative contraindication, and the presence of an unruptured aneurysm >7 mm might also influence this decision. Given these factors and the patient's NIHSS score indicating significant deficit, the decision would likely lean towards caution but could still consider treatment given the early presentation and potential benefits.

The best answer is: $\boxed{Yes}$

22. Llama-4-Scout-17B-16E-Instruct (Structured Prompting) - Case C

**Step 1: Extract Critical Information**

- Patient demographics: Maria Gonzales, 42 years old, female.
- Time of symptom onset: 2 hours prior to evaluation.
- Current time of evaluation: Not specified, but it's 2 hours after symptom onset.
- Neurological deficits: Dysarthria, difficulty walking, left-sided incoordination, NIHSS score of 6.
- Vital signs: Blood pressure 140/80 mmHg.
- Lab values: Not provided, but CT head is negative for hemorrhage.
- Medical history: Small lacunar stroke of the left basal ganglia 2 months ago with full recovery, 8 mm unruptured intracranial aneurysm of the anterior communicating artery.
- Medications: Not specified.
- Menstrual status: Currently menstruating with normal flow.

**2: Time Analysis**

- Time since symptom onset: 2 hours.
- Time window for thrombolytic therapy: Within 0-3 hours or 3-4.5 hours. The patient is within the 0-3 hour window.

**3: Contraindication Check**

- Absolute contraindications:
  - History of intracranial hemorrhage (not present).
  - Known coagulopathy or bleeding diathesis (not mentioned).
  - Recent major surgery or trauma (not mentioned).
  - Recent ischemic stroke within 3 months (occurred 2 months ago).
  - Intracranial neoplasm (not mentioned).
  - Severe uncontrolled hypertension (BP <185/110 mmHg).
  - Suspected subarachnoid hemorrhage (not indicated).
- Relative contraindications:
  - Unruptured intracranial aneurysm (<10 mm).
  - Menstruating with normal flow.

**4: Decision Process**

- The patient is within the 0-3 hour window.
- No absolute contraindications are present.
- The patient has a small unruptured intracranial aneurysm and is menstruating, which are considered relative contraindications.

**5: Guidelines Application**

- Age: Eligible (Class I; Level of Evidence A).
- Stroke severity: Disabling stroke symptoms (NIHSS 6), recommend thrombolysis (Class I; Level of Evidence A).
- Menstruation: Thrombolysis may be considered in women who are menstruating without menorrhagia (Class IIa; Level of Evidence C).
- Unruptured intracranial aneurysm (<10 mm): Administration of thrombolytics is reasonable (Class IIa; Level of Evidence C).

**6: Recommendation**

Given the patient is within the therapeutic time window, has disabling stroke symptoms, and no absolute contraindications, thrombolytic therapy with tPA/alteplase is recommended. The presence of a small unruptured intracranial aneurysm and menstruation are noted but do not contraindicate treatment according to guidelines.

**7: Risk/Benefit Discussion**

"Good morning, Maria. We've discussed your condition, and I want to explain the treatment options. You had a stroke 2 hours ago, and we can offer a medication called alteplase to help dissolve the clot causing your symptoms. This can significantly improve your chances of recovering well. However, like all treatments, it comes with risks, including a small chance of bleeding in the brain or elsewhere. Given your current menstrual flow and small aneurysm, we need to monitor you closely. But for someone with a stroke like yours, the benefits of this treatment likely outweigh the risks. We'll keep a close eye on you and adjust as needed. What do you think about moving forward with this treatment?"

The best answer is Yes.

23. Llama-3.3-70B-Instruct-Turbo (Simple Prompting) - Case C

To determine whether Maria Gonzales should be offered thrombolytics, we need to consider several factors based on her clinical presentation and history.

1. Time of onset: She presented 2 hours after the acute onset of symptoms, which is within the time window for thrombolytic therapy. The standard time window for intravenous thrombolysis with tissue plasminogen activator (tPA) is up to 4.5 hours from the time the patient was last seen well, but the benefit is greatest when administered as soon as possible after symptom onset.
2. Symptom severity: Her NIHSS (National Institutes of Health Stroke Scale) score is 6, indicating a mild to moderate stroke. Thrombolytic therapy can be beneficial for patients with mild to severe strokes, but the decision should be individualized based on the patient's overall clinical condition.
3. History of stroke and recovery: She had a full recovery from a previous small lacunar stroke, which suggests that her brain may have some reserve capacity. However, the history of a previous stroke does not inherently preclude the use of thrombolytics.
4. Presence of an unruptured intracranial aneurysm: The presence of an 8 mm unruptured intracranial aneurysm of the anterior communicating artery is a consideration. The use of thrombolytics in patients with unruptured aneurysms is generally approached with caution due to the theoretical risk of aneurysmal rupture. However, guidelines suggest that the presence of an unruptured aneurysm is not an absolute contraindication to thrombolytic therapy, especially if the aneurysm is not in a location that would be directly affected by the thrombolysis.
5. Menstrual status: Being menstruating with a normal flow is not typically a contraindication for thrombolytic therapy.
6. Blood pressure: Her blood pressure is 140/80 mmHg, which is within the acceptable range for thrombolytic therapy. Guidelines typically recommend that blood pressure should be below 185/110 mmHg before administering tPA.
7. CT head negative for hemorrhage: This is crucial, as thrombolytic therapy is contraindicated in the presence of intracranial hemorrhage.

Considering these factors, Maria Gonzales does not have any absolute contraindications to thrombolytic therapy based on the information provided. The decision to offer thrombolytics should be made by a multidisciplinary team, including a neurologist and possibly a neurosurgeon, considering the potential benefits and risks, including the risk associated with her unruptured aneurysm. The team should discuss the individualized risk-benefit analysis with the patient and her family to make an informed decision.

24. Llama-3.3-70B-Instruct-Turbo (Structured Prompting) - Case C

**Extract Critical Information**

1. Patient Demographics:
   - Age: 42 years
   - Sex: Female
2. Time of Symptom Onset and Evaluation:
   - Time of symptom onset: 2 hours before evaluation
   - Current time of evaluation: Not specified, but 2 hours after symptom onset
3. Neurological Deficits:
   - Symptoms: Dysarthria, difficulty walking, left-sided incoordination
   - NIHSS score: 6
4. Vital Signs:
   - Blood Pressure: 140/80 mmHg
5. Relevant Lab Values:
   - Not specified
6. Imaging Findings:
   - CT head: Negative for hemorrhage
   - Known 8 mm unruptured intracranial aneurysm of the anterior communicating artery
7. Medical History and Medications:
   - History of small lacunar stroke of the left basal ganglia 2 months ago with full recovery
   - No history of intracranial hemorrhage related to the prior stroke
   - No recent major surgery or cancer
   - Menstruating with normal flow

**Time Analysis**

1. Time Since Symptom Onset: 2 hours
2. Applicable Time Window: 0-3 hours

**Contraindication Check**

1. Absolute Contraindications:
   - None explicitly mentioned that cannot be quickly corrected or are relevant to this case (e.g., no intracranial hemorrhage, no recent major surgery, etc.)
2. Relative Contraindications:
   - Known 8 mm unruptured intracranial aneurysm
   - Menstruating with normal flow
   - History of small lacunar stroke 2 months ago
3. Special Circumstances:
   - The presence of an unruptured intracranial aneurysm and menstruation are considerations but are addressed in the guidelines.

**Decision Process**

Given the information:

- The patient is within the 0-3 hour time window.
- There are no absolute contraindications.
- Relative contraindications include the unruptured intracranial aneurysm and menstruation, but guidelines suggest that intravenous alteplase may be reasonable for patients with small or moderate-sized unruptured intracranial aneurysms (Class IIa; Level of Evidence C) and for women who are menstruating without a history of menorrhagia (Class IIa; Level of Evidence C).

**Recommendation**

Yes, thrombolytic therapy with intravenous alteplase can be offered to this patient, considering the benefits likely outweigh the risks in this scenario. The patient's symptoms suggest a disabling stroke, and the time from symptom onset is within the recommended window for treatment. The presence of an unruptured intracranial aneurysm and menstruation are relative contraindications, but based on the guidelines, they do not necessarily preclude treatment.

**Risk/Benefit Discussion**

"Ms. Gonzales, based on your symptoms and our evaluation, you are a candidate for a treatment called intravenous alteplase, which can help dissolve the clot causing your stroke. This treatment is time-sensitive and works best when given promptly. While you have an unruptured intracranial aneurysm and are menstruating, our guidelines suggest that the benefits of this treatment may outweigh the risks for you. However, it's essential to discuss these risks, which include a small chance of bleeding in the brain or worsening of the aneurysm, although the data suggests that the risk of significant bleeding with an aneurysm of this size is low. We also need to monitor your menstrual bleeding after the treatment. Given your current condition and the potential for significant improvement with this treatment, we recommend proceeding with intravenous alteplase. Do you have any questions or concerns about this treatment?"

25. r1-1776 (Simple Prompting) - Case A

Recommendation for Thrombolytics in Tina Patel's Case:

1. Eligibility Assessment:
   - Time Window: Presentation at 75 minutes post-onset is within the 4.5-hour window for alteplase.
   - NIHSS Score: 9 (moderate severity), supporting potential benefit from thrombolysis.
   - CT Findings: No hemorrhage or frank hypodensity, ruling out contraindications for thrombolytics.
   - Medical History: No major bleeding, surgery, or cancer, which minimizes contraindications.
2. Critical Issue: Blood Pressure (BP):
   - Current BP: 190/115 mmHg exceeds the threshold of 185/110 mmHg for safe thrombolytic administration.
   - Guideline Requirement: BP must be reduced to <185/110 mmHg before initiating thrombolytics to mitigate hemorrhage risk.
3. Decision:
   - Immediate Action: Aggressive BP management with antihypertensives (e.g., IV labetalol or nicardipine) is required.
   - If BP is Controlled: Offer thrombolytics once BP is sustained below 185/110 mmHg.
   - If BP Remains Elevated: Thrombolytics are contraindicated due to increased risk of complications.

Conclusion: Tina Patel should be offered thrombolytics only if her blood pressure is successfully reduced to <185/110 mmHg. If this cannot be achieved, thrombolysis should not be administered. The emergency team must prioritize rapid BP control to determine eligibility.

26. r1-1776 (Structured Prompting) - Case A

Recommendation:
Yes, thrombolytic therapy (tPA/alteplase) should be offered after blood pressure is controlled to <185/110 mmHg.

### Supporting Rationale

### Time Analysis

- Symptom onset to evaluation: 75 minutes (well within the 0–3-hour window).
- NIHSS Score 9 (moderate, disabling deficits).

### Absolute Contraindications Check

1. Imaging: CT head shows no hemorrhage or frank hypodensity (no exclusion).
2. Blood Pressure (BP): Current BP is 190/115 mmHg (absolute contraindication if not controlled to <185/110 mmHg).
   - Guideline: BP must be stabilized <185/110 mmHg before tPA (Class I; Level B).
3. No other absolute contraindications (no recent surgery, trauma, coagulopathy, or hemorrhage risk factors).

### Relative Contraindications

- Hypertension: Requires urgent management but is modifiable.

### Decision Process

- The patient is within the 0–3-hour window with disabling deficits (NIHSS 9).
- BP is the sole barrier. Per guidelines, tPA is recommended if BP can be safely lowered.
- Given her history of well-controlled hypertension, aggressive antihypertensive therapy (e.g., IV labetalol, nicardipine) should be initiated to achieve BP <185/110 mmHg.

### Risk/Benefit Discussion for the Patient

*"Ms. Patel, you’re experiencing a stroke caused by a blockage in a brain artery. We recommend a clot-busting medication called alteplase (tPA), which can improve your chances of recovering arm strength and facial movement. However, your blood pressure is currently too high, which increases the risk of bleeding in the brain if we give the medication.

Benefits:

- 30% higher chance of minimal or no disability at 3 months.
- Improved recovery of right arm weakness and facial droop.

Risks:

- 6% risk of symptomatic brain hemorrhage (higher if BP remains elevated).
- 2% risk of major systemic bleeding.

Next Steps:

- We’ll immediately administer medications to lower your blood pressure.
- If we can stabilize it below 185/110 within the next 15–20 minutes, we strongly advise proceeding with tPA.
- If your BP remains elevated, the risks outweigh the benefits, and we’ll pursue alternative treatments like blood pressure control and close monitoring."*

### Alternative Management if BP Uncontrolled

- Blood pressure control: Target <180/105 mmHg with IV antihypertensives.
- Aspirin: Initiate 325 mg orally/rectally if no hemorrhage on repeat imaging.
- Neuroprotective measures: Optimize oxygenation, glucose control, and avoid hyperthermia.
- Secondary prevention: Statin, antihypertensive regimen, and vascular workup (e.g., carotid ultrasound, echocardiogram).

Conclusion: tPA is appropriate if BP is controlled per guidelines. Immediate action to lower BP is critical to eligibility.

27. r1-1776 (Simple Prompting) - Case B

Based on the presented case and current guidelines, Joseph Mercado should not be offered thrombolytics (e.g., alteplase). Here's the critical analysis:

### Key Contraindications:

1. INR >1.7 on Warfarin:
   - His INR is 1.8, exceeding the threshold of ≤1.7 for thrombolytic eligibility in patients on warfarin. This elevates the risk of hemorrhagic complications, including intracranial hemorrhage, which is a strict contraindication per AHA/ASA guidelines.

### Other Considerations:

- Time Window: He is within the 4.5-hour window for thrombolysis (last known normal 3 hours ago).
- Blood Pressure: Systolic BP 175 mmHg is below the 185 mmHg cutoff and manageable.
- NIHSS Score: A high score (28) indicates severe stroke but is not a contraindication.
- Imaging: CT shows early ischemic changes without hemorrhage or significant hypodensity, which does not exclude thrombolysis.
- Basal Cell Carcinoma Excision: Minor surgery does not qualify as a contraindication (major surgery/trauma within 14 days is required for exclusion).

### Conclusion:

The elevated INR (1.8) while on warfarin is an absolute contraindication for thrombolysis. Alternative interventions, such as mechanical thrombectomy (if imaging confirms a large vessel occlusion), should be considered if appropriate. However, thrombolytics are not safe in this case due to the bleeding risk associated with his supratherapeutic INR.

Final Recommendation: Withhold thrombolytics. Pursue further evaluation for possible endovascular therapy and optimize anticoagulation management.

28. r1-1776 (Structured Prompting) - Case B

Recommendation:
No, thrombolytic therapy (tPA/alteplase) should not be offered.

### Supporting Rationale

1. Absolute Contraindication (INR >1.7):
   - The patient is on warfarin with an INR of 1.8. Per guidelines:
     - *"Intravenous alteplase in patients who have a history of warfarin use and an INR >1.7 is not recommended"* (Class III; Level of Evidence B).
   - This elevates the risk of symptomatic intracranial hemorrhage (sICH) and is an absolute exclusion criterion.
2. Time Window:
   - Symptom onset was 3 hours ago, placing him within the 0–3-hour window. However, the elevated INR supersedes time eligibility.
3. Other Considerations:
   - Blood Pressure (175/90 mmHg): Within acceptable range (<185/110 mmHg) and can be stabilized before administration, but irrelevant due to INR contraindication.
   - NIHSS 28 (Severe Stroke): While high NIHSS increases sICH risk, guidelines do not exclude treatment for severity alone in the 0–3-hour window.
   - Basal Cell Carcinoma Excision (1 week ago): Minor surgery is not a contraindication (guidelines reference major surgery within 14 days as a relative concern).

### Alternative Management

- Mechanical Thrombectomy: If imaging confirms a large vessel occlusion (likely given NIHSS 28), endovascular therapy should be urgently considered.
- Supportive Care: Blood pressure management, glycemic control, and monitoring for complications.

### Risk/Benefit Discussion to Patient/Family

*"Mr. Mercado has suffered a severe stroke, and we understand the urgency to act. However, his blood tests show that his blood is thinner than the safe limit for clot-busting medication (tPA). Giving tPA in this situation could risk serious bleeding in the brain, which would be harmful. While this treatment isn’t an option, we will immediately evaluate him for a procedure to remove the clot mechanically if possible. Our team will prioritize stabilizing his condition and preventing further damage."*

Guidelines Adhered To: INR exclusion for warfarin-treated patients (Class III; Level B) and time-window criteria (Class I; Level A).

29. r1-1776 (Simple Prompting) - Case C

Answer:
Maria Gonzales should not be offered thrombolytics (e.g., alteplase) for the following reasons:

1. Prior Ischemic Stroke Within 3 Months:
   She had a lacunar stroke 2 months ago. Current guidelines (AHA/ASA) contraindicate thrombolysis in patients with any prior stroke (ischemic or hemorrhagic) within the preceding 3 months, regardless of recovery. This is due to an increased risk of hemorrhage or recurrent ischemic complications.
2. Unruptured Intracranial Aneurysm:
   Her 8 mm anterior communicating artery aneurysm is a relative contraindication. While not an absolute exclusion, aneurysms ≥7 mm in high-risk locations (e.g., anterior communicating artery) carry a higher rupture risk, and thrombolytics may theoretically exacerbate this risk. Shared decision-making is advised, but most clinicians would avoid tPA in this scenario.

Additional Considerations:

- NIHSS Score 6: While her symptoms are moderate, thrombolysis is still indicated for disabling deficits if no contraindications exist.
- Normal Menstruation: Menstruation is not a contraindication unless accompanied by significant bleeding (not present here).
- BP 140/80 mmHg: Well within acceptable limits for thrombolysis.
- CT Negative for Hemorrhage: Rules out intracranial hemorrhage but does not override other contraindications.

Conclusion: The combination of her recent stroke and intracranial aneurysm makes thrombolytics inadvisable. Alternative therapies (e.g., antiplatelets, supportive care) and urgent vascular imaging (e.g., CTA/MRA) to assess for large-vessel occlusion and potential mechanical thrombectomy should be prioritized.

30. r1-1776 (Structured Prompting) - Case C

Recommendation:
Requires shared decision making with modified risk/benefit discussion

### Supporting Rationale

1. Time Window:
   - Symptom onset: 2 hours prior (within the 0–3-hour window). Eligible for alteplase by time criteria (Class I; Level A).
2. Absolute Contraindications:
   - None identified:
     - CT head negative for hemorrhage.
     - No history of intracranial hemorrhage or coagulopathy.
     - Blood pressure (140/80 mmHg) <185/110 mmHg.
3. Relative Contraindications/Key Considerations:
   - Prior Ischemic Stroke within 3 Months:
     - Maria had a lacunar stroke 2 months ago with full recovery.
     - Guideline: Prior ischemic stroke within 3 months is a relative contraindication ("may be harmful," Class III; Level B). However, expert consensus suggests thrombolysis may be considered in selected cases (e.g., small infarct, good recovery, >1 month since prior stroke).
   - Unruptured Intracranial Aneurysm (8 mm):
     - Not a contraindication. Small/moderate (<10 mm) unruptured aneurysms allow thrombolysis (Class IIa; Level C).
   - Menstruation:
     - Not a contraindication. Increased menstrual flow is possible but manageable (Class IIa; Level C).
4. Benefit-Risk Assessment:
   - Potential Benefits: Improved odds of functional recovery (NIHSS 6 with disabling left-sided incoordination/dysarthria).
   - Risks:
     - Slightly elevated symptomatic intracerebral hemorrhage (sICH) risk due to recent stroke (2 months ago).
     - Aneurysm rupture risk with thrombolysis is very low but not zero.

### Alternative Management

If thrombolysis is declined:

- Antiplatelet therapy (aspirin), statin, blood pressure control, and close monitoring.

### Risk/Benefit Discussion for Provider

Provider Speech:
*"Maria, you’re having a stroke affecting your speech and coordination. The treatment we’re considering, alteplase, can dissolve clots and improve recovery. Benefits include a higher chance of regaining function, but there are risks.*

- *Key Risks:*
  1. *Bleeding in the Brain (1 in 15 patients): Your prior stroke 2 months ago slightly raises this risk, but your recovery was excellent, which is reassuring.*
  2. *Aneurysm Rupture (Very Rare): Your brain scan shows an 8mm bulge in a blood vessel, but studies suggest this size rarely ruptures with treatment.*
  3. *Heavier Menstrual Flow: Expected but manageable.*
- *Benefits: Without treatment, your walking and speech may stay impaired. With alteplase, you have a better chance of improvement.*

*Given your recent stroke, the decision is nuanced. Many experts would still recommend treatment given your young age and disabling symptoms, but we need to weigh this together. What questions or concerns do you have?"*

31. GPT-5.2 Thinking Low (Simple Prompting) - Case A

Yes—she should be offered IV thrombolysis if her blood pressure can be safely lowered to the treatment threshold.

- She is within the IV thrombolysis time window (75 minutes from onset; ≤4.5 hours).

- She has a disabling neurologic deficit (NIHSS 9 with facial droop and arm weakness).

- Noncontrast CT shows no hemorrhage or established large infarct (“no frank hypodensity”).

- Glucose is acceptable (125 mg/dL).

- No major listed contraindications (recent major surgery/bleeding, intracranial hemorrhage, etc.).

The main current barrier is blood pressure:

- IV thrombolysis requires BP <185/110 mmHg before bolus and maintained <180/105 mmHg afterward.

- With BP 190/115, she should receive IV antihypertensive therapy (e.g., labetalol or nicardipine/clevidipine) and, if she reaches <185/110 without excessive delay, proceed with IV alteplase (or tenecteplase per local protocol).

- If BP cannot be controlled to the required threshold, thrombolysis should not be given.

32. GPT-5.2 Thinking Low (Structured Prompting) - Case A

\# 1) Extracted critical information (from vignette only)

| Category | Details |
| --- | --- |
| Demographics | 55-year-old woman |
| Presentation/deficits | Sudden right facial droop + right arm weakness |
| Stroke severity | NIHSS 9 (moderate; typically disabling) |
| Time last known well (symptom onset) | 75 minutes before ED presentation |
| Time of evaluation | At ED arrival (same timepoint given); thrombolysis decision being considered now |
| Glucose | 125 mg/dL |
| Imaging | Noncontrast CT head:No hemorrhage; no frank hypodensity |
| Blood pressure | 190/115 mmHg |
| Relevant history provided | Well-controlled hypertension;No history of major surgery, major bleeding, or any cancer |
| Medications / anticoagulants / labs (INR, platelets, aPTT) | Not provided in vignette |

\# 2) Time analysis

| Item | Result |
| --- | --- |
| Time since symptom onset | ~75 minutes (1 hour 15 minutes) |
| Applicable treatment window | 0–3 hours window |

Guideline tie-in:IV alteplase is recommended for selected patients treatable within 3 hours of onset (Class I; Level of Evidence A).

\# 3) Contraindication check (using only provided guideline criteria)

| Potential issue | Vignette finding | Guideline implication |
| --- | --- | --- |
| Intracranial hemorrhage on CT | No | Not excluded by ICH criterion (ICH would be Class III harm; LOE C) |
| Extensive early ischemic change / frank hypodensity | No frank hypodensity | Alteplase recommended when EICs are mild–moderate “other than frank hypodensity” (Class I; LOE A) |
| Glucose too low/high (mimic concern) | 125 mg/dL | Meets glucose criterion (>50 mg/dL) for alteplase use (Class I; LOE A) |
| Blood pressure above treatment threshold | 190/115 mmHg | Must lower to <185/110 and ensure stability before alteplase; if it can be lowered safely, alteplase is recommended (Class I; LOE B) |
| Major surgery/major trauma/recent bleeding/malignancy (as provided) | None reported | No exclusion based on provided history |
| Coagulopathy/anticoagulants (INR/platelets/aPTT, DOAC use, etc.) | Not provided | Guidelines allow not delaying alteplase for labs if no reason to suspect abnormal tests (Class IIa; LOE B). Vignette gives no suspicion of coagulopathy/anticoagulant use, but it is not explicitly stated. |

\# 4) Decision process

Recommendation: Yes — offer IV alteplase, but only after BP is lowered and confirmed stable below threshold.

Rationale linked to the provided guidelines:

- She is within 0–3 hours (75 minutes) → IV alteplase is recommended for eligible patients (Class I; Level of Evidence A).

- Deficits with NIHSS 9 are consistent with a disabling ischemic stroke → treating disabling strokes within 3 hours is indicated (Class I; Level of Evidence A).

- CT shows no hemorrhage and no frank hypodensity → imaging does not exclude; alteplase is recommended when there is not frank hypodensity (Class I; Level of Evidence A).

- The only clear barrier in the vignette is BP 190/115. Guidelines state alteplase is recommended if BP can be lowered safely to <185/110 and stabilized before starting alteplase (Class I; Level of Evidence B), and then maintained <180/105 for 24 hours (Class I; Level of Evidence B).

Operationally (based on guideline language):

- Give antihypertensives to bring BP below 185/110, confirm it is stable at that level, then proceed with alteplase promptly (time-sensitive benefit; Class I; LOE A for urgency/rapid treatment).

\# 5) If BP cannot be controlled quickly (alternative plan)

If BP cannot be reduced and kept <185/110 in a timely fashion, then do not give IV alteplase (because the “treat if BP can be lowered safely” requirement would not be met; Class I; LOE B implies BP control is necessary).

Alternative management from vignette-supported actions:

- Continue acute stroke care with BP management and close neurologic monitoring.

- Continue ischemic stroke evaluation and management pathways (the vignette/guidelines provided do not give additional specific alternatives beyond thrombolysis eligibility criteria).

\# 6) Risk/benefit discussion script (provider-facing)

“Your symptoms and exam are consistent with an acute ischemic stroke, and you arrived very early—about 75 minutes after symptoms began. A clot-busting medication called alteplase can improve the chance of being independent later if given quickly, and treatment benefit is strongly time-dependent (Class I, Level A). Your CT scan does not show bleeding, which is required before giving this medication, and your blood sugar is in a safe range for treatment (Class I, Level A).

The main issue right now is your blood pressure, which is above the safe limit for giving alteplase. The guideline recommends giving alteplase if we can lower your blood pressure safely to below 185/110 and confirm it stays stable there before starting the medication, and then keep it controlled afterward (Class I, Level B). We can give IV blood pressure medicines now to bring it down quickly and safely.

The major risk of alteplase is bleeding, including bleeding in the brain, which can be serious. The potential benefit is a better chance of recovery and less long-term disability. Because you are early in the window and your stroke symptoms are significant, the potential benefit is meaningful if we can safely control your blood pressure. If we can’t get the blood pressure down and stable soon, then we should not give alteplase and will proceed with other stroke care.”

If you want, share whether she is on any antiplatelet/anticoagulant medication or if any platelet count/INR/aPTT are available, and I can re-check eligibility using only the provided guideline criteria.

33. GPT-5.2 Thinking Low (Simple Prompting) - Case B

No.

IV thrombolysis (alteplase or tenecteplase) is contraindicated in a patient taking warfarin if the INR is >1.7. His INR is 1.8, so he should not be offered IV thrombolytics even though he is within 3 hours of last-known-well, his BP is below 185/110, and his recent basal cell excision would not typically count as “major surgery.”

He should instead be urgently evaluated for large-vessel occlusion and offered endovascular thrombectomy if eligible (given his severe deficit/NIHSS 28 and early CT without hemorrhage).

34. GPT-5.2 Thinking Low (Structured Prompting) - Case B

\# 1) Extract Critical Information (from vignette only)

| Category | Key details |
| --- | --- |
| Demographics | 78-year-old man |
| Presenting syndrome | Aphasia + severe left-sided weakness (disabling) |
| Stroke severity | NIHSS 28 (severe) |
| Last known well (LKW) | 3 hours ago |
| Time of evaluation | Not explicitly stated; presentation implies evaluation occurring at ~3 hours from LKW |
| Blood pressure | 175/90 mmHg |
| Antithrombotic/anticoagulant use | Atrial fibrillation on warfarin |
| Coagulation labs | INR 1.8 |
| Other labs provided | None (no glucose, platelets, aPTT reported) |
| Imaging | CT head: early subtle ischemic changes; no significant hypodensity; no hemorrhage |
| Recent procedures/bleeding | No recent major surgery or major bleeding; basal cell carcinoma excised last week |

\# 2) Time Analysis

| Item | Result |
| --- | --- |
| Time since symptom onset / LKW | ~3 hours |

Applicable guideline window	0–3 hours (borderline at 3 hours based on provided information)

\# 3) Contraindication Check (systematic)

| Potential issue | Vignette finding | Guideline criterion | Implication |
| --- | --- | --- | --- |
| Intracranial hemorrhage on CT | None | Do not give if CT shows acute ICH (Class III; Level of Evidence C) | Not excluded on this basis |
| Extensive frank hypodensity | No (“no significant hypodensity”) | Not recommended if extensive clear hypoattenuation/frank hypodensity (Class III; Level of Evidence A). Recommended if early ischemic changes mild–moderate (Class I; Level of Evidence A) | Imaging supports eligibility |
| Blood pressure too high | 175/90 | tPA recommended if BP can be lowered to <185/110 and stabilized (Class I; Level of Evidence B) | Meets BP criterion as stated |
| Warfarin with elevated INR | Warfarin; INR 1.8 | IV alteplase in warfarin users with INR >1.7 is not recommended (Class III; Level of Evidence B). Safety/efficacy with INR >1.7 not recommended (Class III; Level of Evidence C) | Absolute exclusion based on provided guideline |
| Recent surgery/trauma | Basal cell excised last week; “no recent major surgery” | Major surgery in preceding 14 days: may be considered case-by-case (Class IIb; Level of Evidence C) | Not the deciding factor here given INR exclusion |
| Age | 78 | Within 3 hours: alteplase equally recommended across age groups (Class I; Level of Evidence A) | Age is not an exclusion |
| Stroke severity | NIHSS 28 (severe) | Severe symptoms: alteplase indicated within 3 hours (Class I; Level of Evidence A) | Would favor treatment if not excluded |

\# 4) Decision Process (apply guideline logic)

Recommendation: No — do not offer IV alteplase (tPA).

Reason: The vignette provides a definitive exclusion: warfarin use with INR 1.8 (>1.7). Per the provided guideline, IV alteplase in patients with a history of warfarin use and INR >1.7 is not recommended (Class III; Level of Evidence B).

Although he is within the treatment time window (~3 hours), has severe disabling deficits (NIHSS 28), blood pressure below the treatment threshold, and CT without hemorrhage or extensive frank hypodensity, the INR criterion is a guideline-based contraindication in the supplied reference.

\# 5) If “No”: Appropriate Alternative Management (based on what can be stated from vignette/guideline set)

- Do not administer IV alteplase due to INR >1.7 (Class III; Level of Evidence B).

- Continue acute ischemic stroke management without alteplase, including close neurologic and blood pressure monitoring.

- Reassess for other acute stroke interventions as appropriate to the presenting syndrome (not specified in the provided guidelines excerpt).

\# Risk / Benefit Discussion Script (what the provider can say)

“Based on your CT scan, there is no bleeding in the brain, and your symptoms are severe enough that clot-busting medicine would normally be considered because it can improve the chance of recovery when given early. However, you take warfarin, and your blood test today shows your INR is 1.8. Under our stroke treatment rules, when the INR is above 1.7, giving alteplase is not recommended because the risk of dangerous bleeding in or around the brain is too high. Because of that, we should not give the clot-busting medication. We will instead focus on other stroke treatments and close monitoring while we continue urgent stroke care.”

35. GPT-5.2 Thinking Low (Simple Prompting) - Case C

No—she should not be offered IV thrombolysis (alteplase/tenecteplase) because she had an ischemic stroke 2 months ago. An ischemic stroke within the prior 3 months is an AHA/ASA exclusion criterion for IV thrombolytics due to increased risk of symptomatic intracranial hemorrhage.

Her other features do not by themselves exclude thrombolysis:

- An 8 mm unruptured anterior communicating artery aneurysm is generally not a contraindication (risk concern is higher with larger aneurysms, but small unruptured aneurysms are typically treated as acceptable).

- Normal menstruation is not a contraindication.

- BP 140/80 and CT negative for hemorrhage would otherwise be acceptable.

Next steps: obtain urgent vascular imaging (CTA/MRA) to look for a treatable large-vessel occlusion and consider endovascular therapy if indicated, plus start appropriate antithrombotic/secondary prevention once hemorrhage risk is excluded and per stroke subtype.

36. GPT-5.2 Thinking Low (Structured Prompting) - Case C

\# 1)Extracted critical information (from vignette)

| Demographics | Maria Gonzales,42-year-old woman |
| --- | --- |
| Symptom onset | Acute onset;arrived/evaluated 2 hours after onset |
| Current time window | 0–3 hours window applies (2 hours since onset) |
| Neurologic deficits | Dysarthria,difficulty walking,left-sided incoordination |
| Stroke severity | NIHSS 6 (moderate;potentially disabling) |
| Blood pressure | 140/80 mmHg (<185/110 threshold) |
| Imaging | CT head negative for hemorrhage |
| Bleeding/gynecologic status | Currently menstruating with normal flow;no menorrhagia reported |
| Relevant past history | Small lacunar stroke (L basal ganglia) 2 months ago with full recovery;known 8 mm unruptured anterior communicating artery aneurysm |
| Medications/labs | Not provided (no anticoagulant use stated;no platelets/INR/glucose provided) |

\# 2)Time analysis

- Time since onset:2 hours

- Guideline window met:Eligible by time for IV alteplase “within 3 hours” (Class I;Level of Evidence A)

\# 3)Contraindication / special-situation check (using only provided guideline statements)

- CT shows no ICH → required exclusion satisfied (alteplase should not be given if ICH on CT:Class III;LOE C)

- Blood pressure acceptable (and below treatment threshold) → hypertension not a barrier (Class I;LOE B for treating if can be lowered to <185/110)

- Menstruation with normal flow,no history of menorrhagia reported → alteplase “probably indicated”;warn may increase flow (Class IIa;LOE C) and monitor bleeding 24h if treated (Class I;LOE C)

- Unruptured intracranial aneurysm 8 mm (<10 mm) → alteplase “reasonable and probably recommended” (Class IIa;LOE C)

- Key issue:prior ischemic stroke 2 months ago (<3 months)

    - “Use of intravenous alteplase…with a prior ischemic stroke within 3 months may be harmful”(Class III;Level of Evidence B)

    - Guideline text also notes selected-case treatment has been supported by expert consensus and that the increased sICH risk “is not well established”(Class IIb;LOE B) and that risks “should be discussed…and weighed”(Class I;LOE C)

\# 4)Decision process (risk–benefit based on the provided guideline text)

Because she is within 3 hours and has a moderate,likely disabling stroke (NIHSS 6),multiple guideline elements support treatment (time window Class I;LOE A;disabling symptoms treated within 3h Class I;LOE A;menstruation normal flow Class IIa;LOE C;aneurysm <10 mm Class IIa;LOE C).

However,she has a prior ischemic stroke within the last 3 months,which the guideline lists as “may be harmful”(Class III;LOE B),yet also provides language supporting individualized consideration in selected cases with explicit risk discussion (Class IIb;LOE B;and risk discussion Class I;LOE C).

\# 5)Recommendation

Requires shared decision making with modified risk/benefit discussion.

- Proceeding with alteplase is not straightforward due to the recent ischemic stroke (2 months) flagged as potentially harmful (Class III;LOE B).

- If treatment is pursued despite this,do so only after explicit discussion that this recent-stroke factor may increase hemorrhage risk and that the guideline calls for weighing risks vs benefits (Class I;LOE C),while noting other features are favorable (within 3h;CT no bleed;BP controlled;small unruptured aneurysm and normal menstruation are not prohibitive per guideline Class IIa;LOE C).

\# If “No alteplase” is chosen (alternative management from what can be inferred within guideline scope)

- Do not give IV alteplase;continue acute stroke care with close monitoring and secondary prevention planning.

- Monitor neurologic status closely and manage blood pressure appropriately.

- (From provided guideline text only:no specific non-tPA reperfusion alternative is specified.)

\# Provider risk/benefit discussion script (patient-facing)

“Your symptoms and exam suggest an acute ischemic stroke,and you arrived about 2 hours after it started,which is within the usual time window where the clot-busting medicine alteplase can improve the chance of recovery. Your brain CT does not show bleeding,and your blood pressure is in a safe range. Also,your known small aneurysm (under 10 mm) and having a normal menstrual period are not,by themselves,reasons to withhold this medicine,though it could make your menstrual bleeding heavier and we would monitor that closely for 24 hours.

The main concern is that you had another ischemic stroke about 2 months ago. The guideline notes that giving alteplase within 3 months of a prior ischemic stroke may be harmful because it could raise the risk of bleeding in the brain. Because of that,we need to weigh the potential benefit of improving your stroke deficits against a potentially higher bleeding risk in your specific situation. If you choose treatment,we will monitor you very closely for any signs of bleeding;if you choose not to,we will focus on supportive stroke care and prevention while we continue evaluation.”
